# GWM-HFN: A Novel Gray-White Matter Heterogeneous Fusion Network for Functional Connectomes

**DOI:** 10.1101/2025.03.05.25323471

**Authors:** Kang Hu, Zhuo-Yan Cai, Yun-Er Xu, Yu Chen, Chu-Chu Zheng, Ze-Qiang Linli

## Abstract

Resting-state functional connectivity (FC) studies have predominantly centered on gray matter (GM), largely overlooking the functional contributions of white matter (WM). However, emerging evidence indicates WM blood-oxygen-level-dependent (BOLD) signals actively shape large-scale brain networks. Current methods to integrate WM face limitations, including challenges in assessing global network properties from bipartite GM-WM connections or the nascent stage of unified connectome models. Here, we introduce the Gray-White Matter Heterogeneous Fusion Network (GWM-HFN), a novel framework that defines GM-GM functional links via the covariance of their interaction profiles with WM bundles defined by a standardized atlas. Validated across six independent datasets, GWM-HFN demonstrates fair short-term (ICC ∼0.36) and slight-to-fair long-term (ICC ∼0.20) test-retest reliability, comparable to conventional GM-based FC. GWM-HFN exhibits distinct topological features, including small-worldness and enhanced modular segregation compared to GM-GM networks, capturing over 40% unique variance and an enhanced ability to resolve the unique connectivity patterns that differentiate individuals. Lifespan analyses reveal global linear declines and complex non-linear age effects in GWM-HFN connectivity, with peak connectivity in early adulthood (∼34 years). Clinically, individuals with autism spectrum disorder (ASD) show GWM-HFN-specific hyperconnectivity, which correlates with symptom severity and offers greater sensitivity than GM-GM FC. Furthermore, GWM-HFN connectivity patterns predict individual differences in cognitive performance, notably in language tasks. The GWM-HFN framework provides a robust and more comprehensive approach to understanding WM-mediated neural communication, integrating functional signals across both GM and WM, and offers promising avenues for developing neuroimaging biomarkers for aging and neuropsychiatric disorders.

## 1. Introduction

Over the last twenty years, resting-state functional magnetic resonance imaging (rs-fMRI) has emerged as a pivotal tool for mapping synchronized neural activity across various brain regions, mapping synchronized neural activity, commonly termed functional connectivity (FC)^1,2^. This innovative imaging technique has significantly enriched our understanding of the brain’s functional connectome, which provides a network-level perspective on the interconnections among brain regions. As a result, researchers have gained deeper insights into the large-scale organizational structure of the brain, the trajectories of development and aging, and the alterations associated with neuropsychiatric disorders^3–6^.

Historically, the majority of studies focusing on the functional connectome have centered on gray matter (GM), where it is commonly assumed that neural activity drives blood-oxygen-level-dependent (BOLD) signals. This GM-centric focus has inadvertently led to a relative neglect of white matter (WM) in connectivity analyses, primarily due to its lower BOLD signal amplitude and its traditional classification as a mere structural conduit facilitating communication between GM regions^7–9^. However, a growing body of evidence is challenging this long-held perspective. The successful detection of WM functional signals, despite their lower amplitude compared to GM signals, is largely attributable to recent methodological advancements. These include specialized preprocessing pipelines that, for instance, apply separate spatial smoothing to WM and GM to prevent signal leakage, and avoid regressing out the WM signal as a nuisance variable^10,11^. Such techniques have enabled the reliable characterization of WM functional networks, suggesting that WM plays a more active role in the dynamics of functional networks than previously acknowledged^10,12–16^.

Recent research, aiming to incorporate WM, has explored several avenues, including WM-only networks, GM-WM interactions, and more unified connectomes, each with its own set of advancements and unresolved challenges. The first approach investigates WM-only functional networks, treating WM parcels as independent nodes and analyzing their intrinsic functional interactions. These studies have demonstrated that WM possesses unique functional connectivity patterns, establishing stable networks that are comparable to those found in GM^13^. Moreover, alterations in WM functional connectivity have been identified in various neurological and psychiatric disorders, such as schizophrenia^17^ and nicotine dependence^18^. However, analyzing WM in isolation may obscure its broader contributions to the coordination of large-scale brain communication. To address this limitation, researchers have introduced a GM-WM network approach, which explicitly models the functional interactions between GM and WM nodes, each defined by distinct atlases^14,19^. This paradigm has revealed significant functional disruptions in psychiatric populations^20,21^, while also providing new insights into how WM contributes to the dynamics of GM networks^22^. Nevertheless, these networks remain structurally constrained, as they primarily capture GM-to-WM connections and lack closed-loop interactions^23^. This structural limitation complicates the application of conventional network analyses, such as assessments of global efficiency or modularity. Additionally, differences in signal amplitude and dynamic properties between WM-WM and GM-GM connections can lead to inconsistencies^12^ when analyzed within a unified framework, ultimately affecting the biological interpretation of network topology.

To overcome these challenges, a third approach—the unified functional connectome—has emerged, aiming to integrate both GM and WM connectivity into a cohesive framework that can capture closed-loop interactions and elucidate comprehensive mechanisms of network integration^24–26^. However, this area of research is still in its infancy, lacking a standardized modeling paradigm. Several promising methods have been put forth, each presenting unique advantages while grappling with methodological hurdles. For instance, Gao et al. introduced a projection-based mapping strategy that mathematically transforms GM-WM functional connections into an equivalent GM-only network, termed the "WM-mediated GM functional connectivity network." By calculating its global efficiency, they discovered a negative correlation between network efficiency and age^24^ as well as a reduction in efficiency among individuals with preclinical Alzheimer’s disease^27^. This innovative approach partially addresses the absence of triangular pathways in bipartite GM-WM networks and highlights distinct topological properties. While this innovative approach enables the calculation of global properties like efficiency, the resulting network is directed and non-reciprocal (i.e., the connectivity from region A to B is not equal to that from B to A), which creates considerable interpretational ambiguity from both methodological and neurobiological standpoints. Another avenue of research has focused on the modulation of GM network organization by WM. Wang et al. developed an edge-centric network analysis approach that illustrates how WM signals significantly regulate GM functional organization, with evidence suggesting that this modulation is impaired in disease populations^26^. Although this elegantly quantifies WM’s modulatory influence, it does not produce an explicit, integrated connectome that can be subjected to further network analysis. More recently, Zu et al. proposed a three-way correlation model that treats WM BOLD signals as a third component within traditional GM functional networks, thereby providing a more comprehensive representation of WM’s mediation of GM interactions^25^. While this offers a rich depiction of the underlying architecture, its three-dimensional structure is incompatible with the vast majority of standard graph-theoretical toolboxes, which require a 2D adjacency matrix, and thus does not resolve the fundamental challenge of analyzing network topology in GM-WM systems; moreover, its mathematical complexity may pose a barrier to broader clinical application. As research in this area continues to evolve, the integration of GM and WM connectivity promises to enhance our understanding of the brain’s complex functional dynamics, ultimately paving the way for improved insights into neuropsychiatric conditions and their underlying mechanisms.

To address these distinct limitations, we propose the Gray-White Matter Heterogeneous Fusion Network (GWM-HFN). This novel framework constructs a unified and weighted GM-GM connectome by defining the functional connectivity between any two GM regions as the covariance of their respective interaction profiles with all WM bundles. This approach uniquely produces a symmetric, intuitive, and fully analyzable 2D network matrix that retains critical information about WM-mediated communication, thereby bridging the gap between descriptive models of WM influence and the practical need for a topologically coherent connectome. To validate our framework, our study rigorously examines the test-retest reliability and topological organization of the GWM-HFN. We juxtapose our findings against those derived from conventional GM-GM connectivity analyses, providing a comprehensive comparison that highlights the unique contributions of our model. Furthermore, we delve into the practical implications of GWM-HFN by investigating age-related changes in connectivity patterns, assessing its clinical relevance, and exploring its correlations with various cognitive and behavioral measures. Through these multifaceted analyses, we aspire to establish a robust and comprehensive framework that not only enhances our understanding of the brain’s functional architecture but also effectively bridges the existing divide between traditional GM-centric methodologies and the active, communicative role of white matter in inter-regional connectivity.

## 2. Results

### 2.1 General Description of Analytical Methods, Datasets and Research Questions

In this research, we presented GWM-HFN, a network framework aimed at merging functional data from two heterogeneous brain structures (GM and WM) by deliberately integrating WM signals into rs-fMRI connectivity evaluations. **Figure 1** illustrates a schematic representation of this method. In summary, following preprocessing, time-series signals were gathered from 90 GM regions (Anatomical Automatic Labeling, AAL atlas; refer to Table S1 for specifics) and 48 WM bundles (JHU-ICBM 48 atlas; refer to Table S2 for specifics). Subsequently, we developed a bipartite GM-WM correlation matrix, where each row represents the functional connectivity profile of a GM region with various WM bundles. Considering that WM acts as the main pathway for long-distance communication between GM regions, we also calculated the covariance of GM-WM connectivity profiles, converting the bipartite matrix into a GM-centered representation that indirectly reflects WM-mediated interactions. The resulting GWM-HFN encapsulates the functional communication strength among GM regions, which is informed by the covariance of their interaction profiles with the brain’s WM bundles. This modification retains WM-derived functional information within a GM-aligned network framework, allowing for the exploration of both global and local network characteristics while incorporating the impact of WM functional connectivity profiles on brain activity.

**Figure 1.**
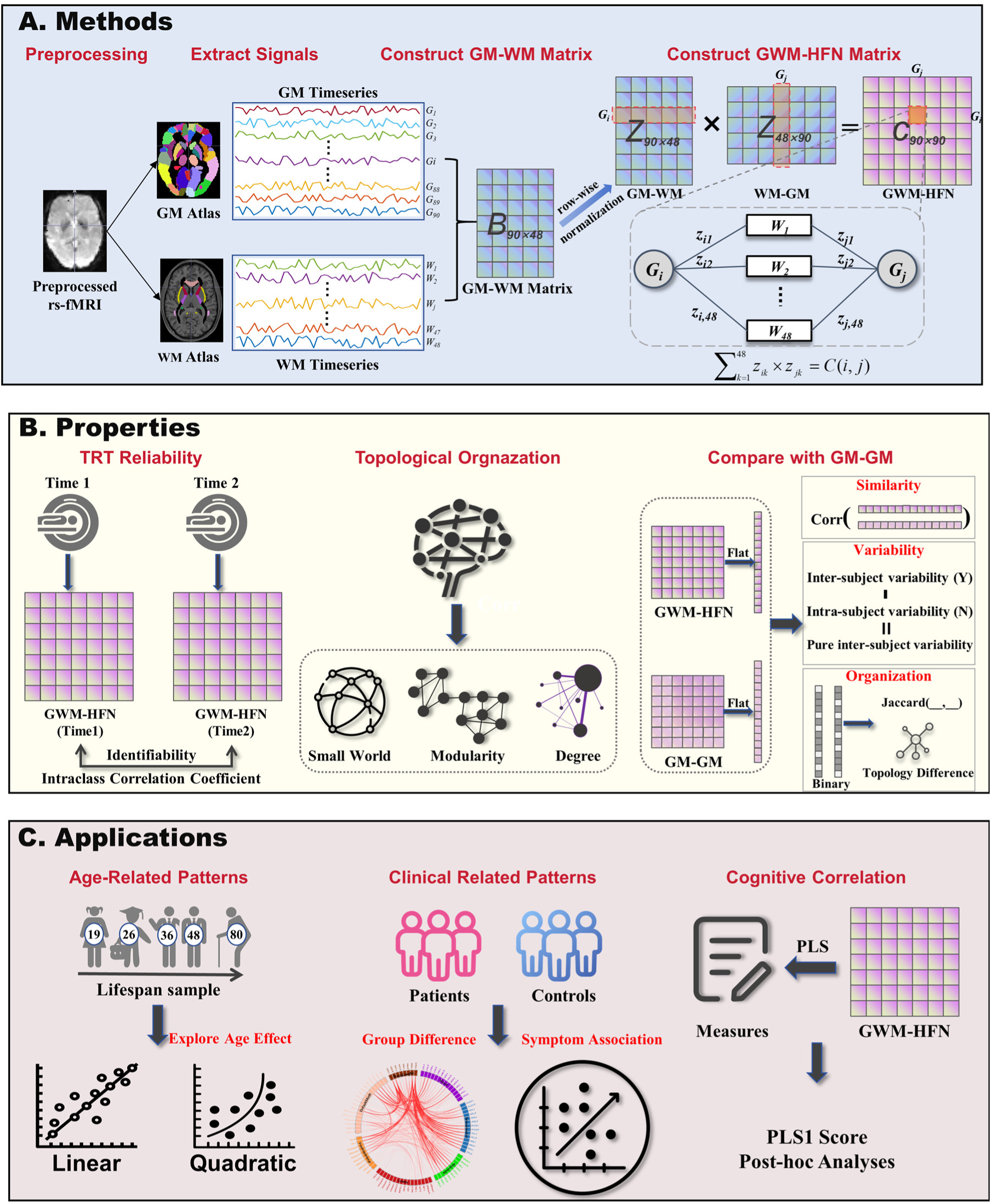
Schematic Overview of the Study Framework. (A) Development of the gray-white matter heterogeneous fusion network (GWM-HFN). This phase initiated with the extraction of gray matter (GM) and white matter (WM) signals from preprocessed rs-fMRI datasets to create a GM-WM connectivity matrix *B*. Following this, the values were standardized row-wise to generate a *Z*-score matrix, highlighting the relative interaction profile of each region within its connectivity framework. Ultimately, the covariance matrix *C*, which was obtained from *Z*, functioned as a GM FC matrix mediated by WM, also referred to as the GWM-HFN. (B) Analysis of GWM-HFN Characteristics. The research evaluated the test-retest reliability of the GWM-HFN network alongside its topological characteristics using graph theoretical metrics. Additionally, comparative assessments between GWM-HFN and conventional GM-GM connectivity networks were carried out. (C) Utilization of GWM-HFN. The practical relevance of the GWM-HFN was investigated across three areas. First, examining its age-related trends, which included both linear and quadratic effects; second, evaluating its use in clinical settings; and third, applying partial least squares (PLS) regression to assess its predictive ability for cognitive and behavioral outcomes.

We meticulously assessed GWM-HFN through various independent datasets to evaluate its dependability, network characteristics, and functional significance, which encompassed test-retest reliability (TRT), structural organization, comparisons to GM-GM connectivity, age-related variations, clinical significance, and cognitive/behavioral associations. In particular, the Southwest University Longitudinal Imaging Multimodal (SLIM) dataset (http://dx.doi.org/10.15387/fcp_indi.retro.slim) was utilized to analyze network patterns, structural organization, and long-term reliability, while the Beijing Normal University (BNU-3) (http://dx.doi.org/10.15387/fcp_indi.corr.bnu3) offered measurements for short-term reliability. The Southwest University Adult Lifespan (SALD) dataset (https://fcon_1000.projects.nitrc.org/indi/retro/sald.html) was employed to examine age-related trajectories of GWM-HFN connectivity within a cross-sectional population. To investigate clinical relevance, we focused on disorder-specific changes in GWM-HFN using the Autism Brain Imaging Data Exchange II (ABIDE-II) project (https://fcon_1000.projects.nitrc.org/indi/abide/abide_II.html), with an emphasis on autism spectrum disorder (ASD). Lastly, the Brain Genomics Superstruct Project (BGSP) dataset (https://www.neuroinfo.org/gsp/) facilitated partial least squares (PLS) regression to connect GWM-HFN connectivity with cognitive and behavioral metrics. A comprehensive description of these datasets is available in the Supporting Information (the “Study Datasets” section), with Table S3 providing a summary of essential demographic details.

To facilitate cross-network comparisons and interpretability, GM nodes were organized into seven functional networks. This was achieved by adapting the Yeo-7 cortical parcellation using a "winner-takes-all" mapping, followed by adjustments based on established anatomical and functional literature. The final seven networks include the Visual Network (VN), Sensorimotor Network (SMN), Attention Network (AN), Limbic Network (LN), Frontoparietal Network (FPN), Default Mode Network (DMN), and Basal Ganglia Network (BGN) (Figure 2A, Table S1). This categorization facilitated a more structured evaluation of GWM-HFN connectivity trends among functionally diverse systems. Additional information regarding node assignments and the relevant methods can be accessed in the Supporting Information (the “Mapping AAL Atlas to Macroscopic Networks” section).

**Figure 2.**
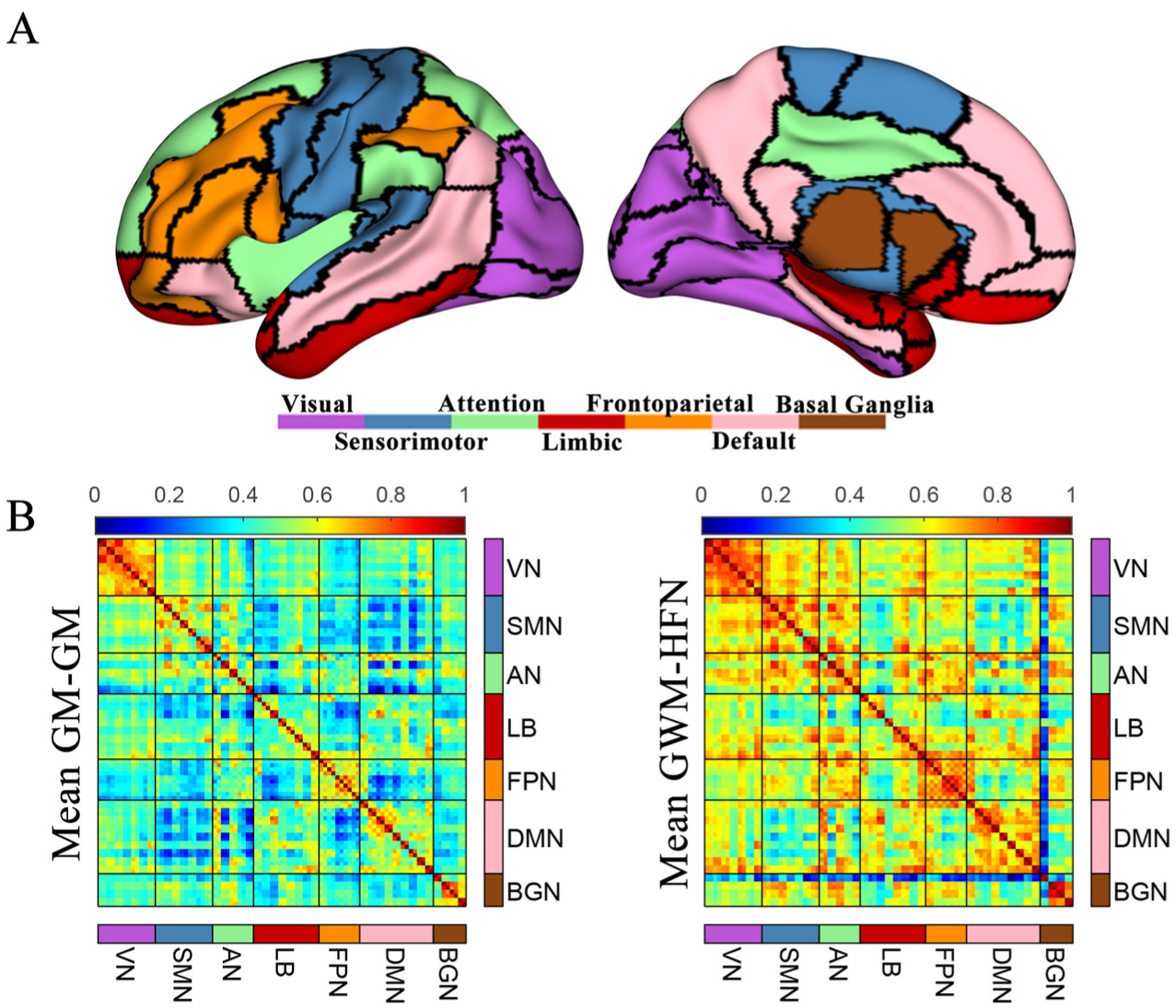
**(A)** A schematic representation of the division of 90 brain regions into seven large-scale networks; **(B)** Group-level mean GM-GM connectivity strength matrix (r-value matrix) and GWM-HFN connectivity strength matrix, both normalized to a range of 0–1 for comparability.

### 2.2 Gray-White Matter Heterogeneous Fusion Network Patterns (SLIM Dataset)

For every one of the 570 younger healthy individuals (ages 17-27) from the SLIM dataset (baseline), we developed GWM-HFN and traditional GM-GM networks. Figure 2B illustrates the mean connectivity matrices at the group level for the constructed GWM-HFN and traditional GM-GM networks. Generally, the connectivity matrices revealed consistently higher intra-network connectivity among large-scale functional subnetworks (such as visual and frontal-parietal networks) and moderately strong inter-network connections, regardless of whether GWM-HFN or GM-GM was analyzed. A visual inspection of the group-average connectivity matrices suggests that the GWM-HFN framework captures connections—both within and between networks—that appear stronger in magnitude compared to those in the GM-GM framework. Rigorous statistical comparisons of network properties derived from these matrices are detailed in Section 2.5. This suggests a fundamental difference in the network topology captured by the two methods.

### 2.3. Multi-level Test-Retest Reliability Assessment (BNU-3 and SLIM Datasets)

The BNU-3 dataset was used to assess short-term test-retest (TRT) reliability, with each participant undergoing three scanning sessions within a single day. In contrast, the SLIM dataset was employed to evaluate long-term TRT reliability. Participants in this dataset underwent up to three scanning sessions, although only a subset completed the second and third sessions. For the purposes of this study, only the first two time points—separated by an average interval of 304.14 days—were used to assess long-term TRT reliability. To provide a comprehensive evaluation and a direct comparison with conventional methods, we assessed reliability using three distinct approaches: (i) pattern similarity via Pearson correlation, (ii) conventional edge-wise reliability via the Intraclass Correlation Coefficient (ICC), and (iii) a more advanced connectome-level identifiability framework.

#### 2.3.1. Pattern Similarity using Pearson Correlation

As illustrated in Figures 3A and 3B, the GWM-HFN exhibited stable and consistent connectivity patterns across time points at the group level in both datasets. Group-averaged connectivity networks showed strong correlations across sessions, with a mean Pearson correlation of r=0.955 for short-term reliability (BNU-3) and r=0.897 for long-term reliability (SLIM). However, this high group-level consistency masks considerable variance at the individual level, where correlations were more modest. In the SLIM dataset, the average individual-level correlation was 0.37 (±0.13), as shown in the lower-right corner of Figure 3A. Similarity, the BNU-3 dataset, the average correlation across time points was approximately 0.53 (*r*_1, 2_ = 0.57; *r*_1, 3_ = 0.52; *r*_2, 3_ = 0.50), as shown in the lower-right corner of Figure 3B. This discrepancy highlights the necessity of individual-level reliability analyses, as high group-level stability does not guarantee that individual-specific connectivity patterns are reliably captured.

**Figure 3.**
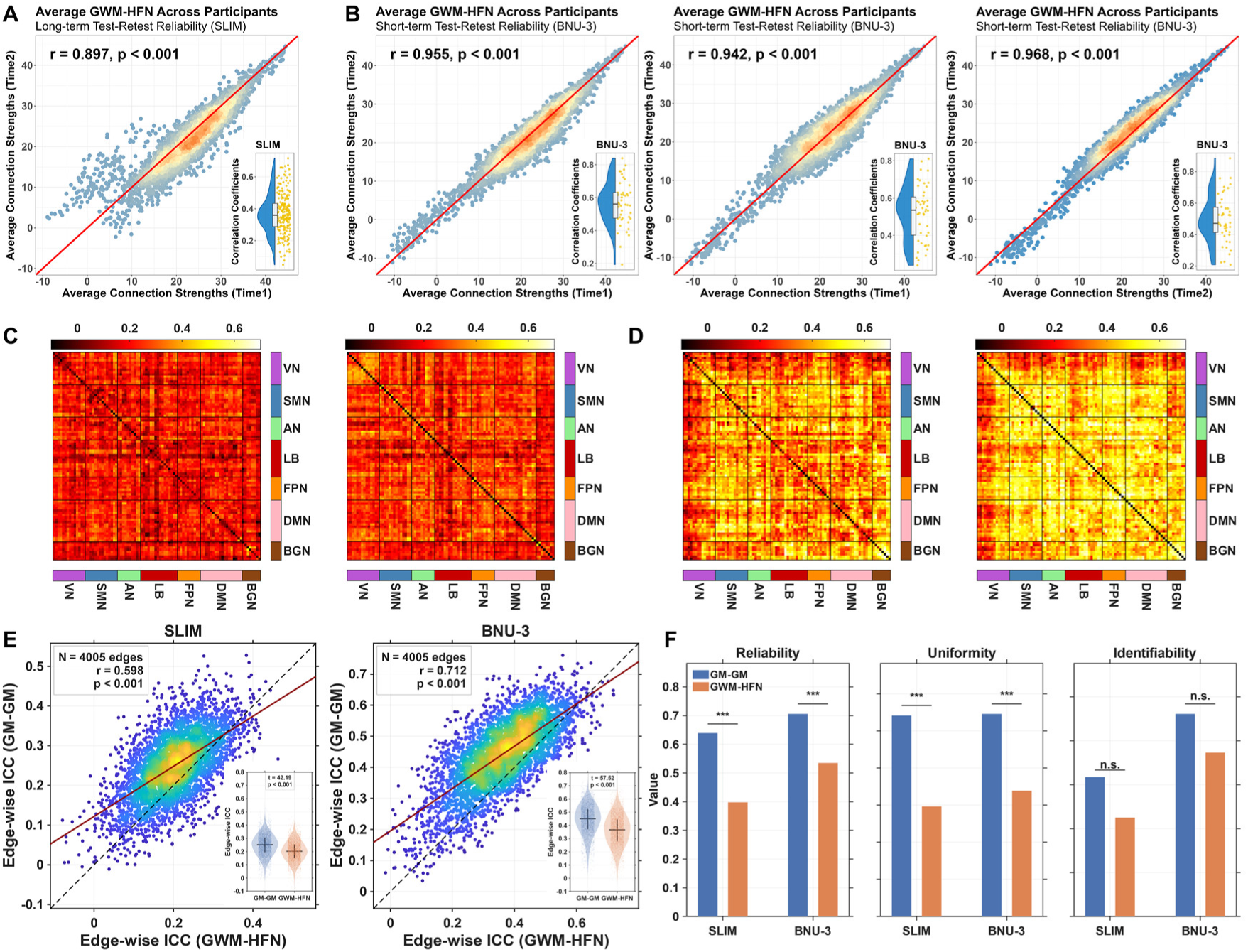
Comprehensive Multi-level and Multi-method Assessment of Test-Retest Reliability. (A, B) Group-level pattern stability versus individual-level similarity for the GWM-HFN. The main scatterplots show the high correlation between group-averaged GWM-HFN connectivity strengths across two sessions for the long-term (A, SLIM dataset) and short-term (B, BNU-3 dataset). The red line indicates the identity line (y=x), and dot color indicates the density of overlapping points. The inset raincloud plot in each panel illustrates the more modest distribution of Pearson correlation coefficients calculated at the individual subject level. (C, D) Edge-wise Intraclass Correlation Coefficient (ICC) matrices. Heatmaps display the ICC value for every connection for both the GWM-HFN (left) and the conventional GM-GM (right) network. Results are shown for the long-term (C, SLIM dataset) and short-term (D, BNU-3 dataset) reliability assessments, with brighter colors indicating higher reliability. (E) Direct comparison of edge-wise ICC values between methods. Scatterplots compare the ICC of each edge from the GWM-HFN (y-axis) against the GM-GM network (x-axis) for the SLIM (left) and BNU-3 (right) datasets. The dashed black line is the identity line, and the red line is the linear fit. The inset violin plots provide a summary comparison of the ICC distributions, confirming the significantly higher mean ICC for the GM-GM method in both datasets. (F) Connectome-level assessment using the identifiability framework. Grouped bar plots compare the GWM-HFN and GM-GM networks on three key metrics: mean intra-individual similarity (Reliability, µ_intra_), mean inter-individual similarity (Uniformity, µ_inter_), and the summary Identifiability Score. While GM-GM shows higher Reliability, GWM-HFN shows significantly lower Uniformity, indicating greater differentiation between individuals. Asterisks denote statistically significant differences.

#### 2.3.2. Edge-wise Reliability using Intraclass Correlation Coefficient (ICC)

We quantified the TRT reliability of individual connections using the ICC. To benchmark our method, we compared the edge-wise ICC of GWM-HFN with that of the conventional GM-GM network (Figure 3C&D). First, we observed a significant positive correlation between the edge-wise ICC values from the two methods across both datasets (see Figure 3E), suggesting a shared spatial pattern of reliability. Following this, as shown in Figure 3E, the GM-GM network exhibited a significantly higher mean edge-wise ICC in both short-term (GM-GM: 0.44 ± 0.11; GWM-HFN: 0.36 ± 0.12; t(4004) = 57.52, p < 0.001) and long-term datasets (GM-GM: 0.24 ± 0.08; GWM-HFN: 0.20 ± 0.08; t(4004) = 42.19, p < 0.001). These results indicate that conventional GM-GM connectivity is more stable at the single-edge level, likely reflecting its sensitivity to strong, generic signals from primary sensory-motor regions that are highly consistent within and across individuals. Notably, and consistent with previous observations, long-term reliability was generally lower than short-term reliability, likely due to increased variability introduced by longer intervals between measurements (e.g., physiological changes and day-to-day fluctuations) ^28,29^.

#### 2.3.3. Connectome-level Assessment using the Identifiability Framework

To provide a more comprehensive assessment of individual-level stability and uniqueness, as suggested by recent literature^30^, we implemented an identifiability framework, which evaluates a connectome’s stability (Reliability, µ_intra_), its distinctiveness from others (Uniformity, µ_inter_), and its overall utility for individual identification (Identifiability).This analysis revealed a critical trade-off between stability and individual specificity (Figure 3F).

Consistent with the ICC results, the GM-GM network showed higher Reliability (mean intra-individual similarity) in both long-term (GM-GM: 0.64 vs. GWM-HFN: 0.40; t(569) = 44.26, p < 0.001) and short-term datasets (GM-GM: 0.71 vs. GWM-HFN: 0.53; t(45) = 21.86, p < 0.001). However, the GWM-HFN demonstrated a striking advantage in Uniformity, exhibiting significantly lower mean inter-individual similarity (µ_inter_) in both long-term (GM-GM: 0.54 vs. GWM-HFN: 0.30; t(569) = 490.77, p < 0.001) and short-term datasets (GM-GM: 0.55 vs. GWM-HFN: 0.34; t(45) = 129.74, p < 0.001). This indicates that while GM-GM networks are more stable on average, GWM-HFN networks are significantly better at capturing the unique connectivity patterns that differentiate one individual from another. Finally, we computed the overall Identifiability scores for both methods. While the GM-GM network yielded numerically higher identifiability scores, these differences were not statistically significant in either the long-term (GM-GM: 1.59 vs. GWM-HFN: 1.12; p = 0.732, bootstrap test) or short-term datasets (GM-GM: 2.30 vs. GWM-HFN: 1.86; p = 1.000, bootstrap test). Taken together, these results reveal a nuanced reliability profile, which will be interpreted in the Discussion.

### 2.4. Topological Organization of the GWM-HFN (SLIM dataset)

For the GWM-HFNs derived from the 570 younger healthy participants from the SLIM time1 dataset, we calculated a set of graph-based network measures across varying sparsity thresholds (0.10-0.34, step=0.01) to study their topological organization, including small-world organization, modularity architecture, degree distribution, and hubs.

#### 2.4.1. Small-World Properties and Modularity

The GWM-HFN consistently demonstrated characteristics indicative of a classic small-world network configuration. This was evidenced by normalized clustering coefficients exceeding 1, alongside characteristic path lengths approximating 1, as illustrated in Figure 4A. Such properties suggest a network topology that facilitates efficient information transfer and integration among disparate brain regions. Moreover, the modularity coefficient (Q) consistently remained above 0.3 across a wide threshold range (Figure 4B), suggesting that explicit WM inclusion produces well-segregated community structures that may capture local clustering mechanisms.

**Figure 4.**
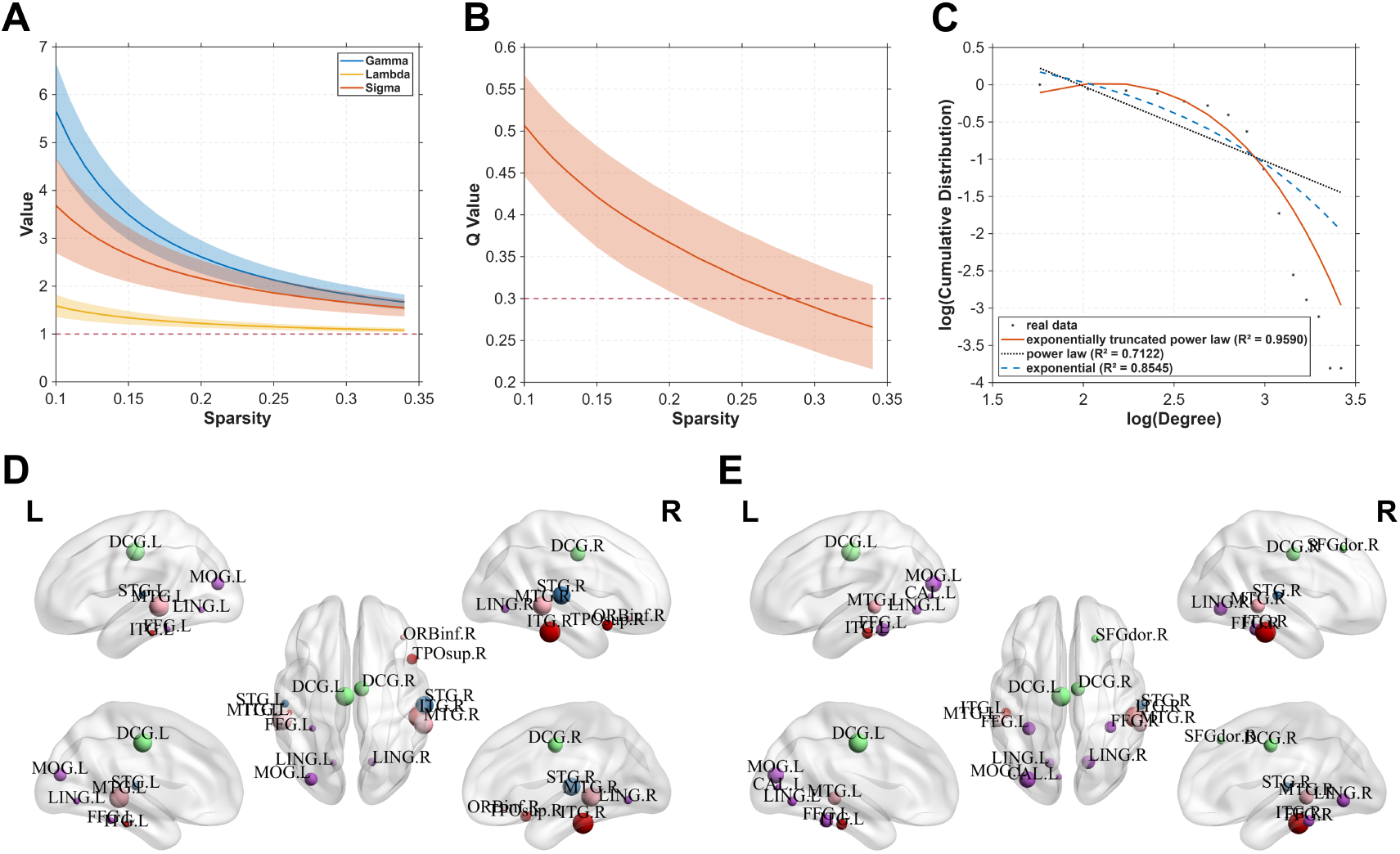
Topological Characterization of the Gray-White Matter Heterogeneous Fusion Network (GWM-HFN). (A) Small-world properties—sigma (σ), normalized clustering coefficient (γ), and normalized characteristic path length (λ)—of the GWM-HFN across a range of sparsity thresholds, presented as mean ± standard deviation. (B) Modularity coefficient (Q) of the GWM-HFN across varying sparsity thresholds, also shown as mean ± standard deviation, indicating robust modular organization. (C) Degree distribution of the group-averaged GWM-HFN, best fitted by an exponentially truncated power-law model, revealing the existence of highly connected brain regions. (D) Identification of 14 hub nodes within the group-averaged GWM-HFN, highlighting their central roles in the brain’s functional connectivity architecture. (E) Hub nodes identified in the benchmark group-averaged GM-GM network, allowing for a direct comparison of the central network architecture revealed by each framework.

#### 2.4.2. Hub Distribution and Degree

The degree distribution observed in the group-level mean GWM-HFN was best represented by an exponentially truncated power law model, achieving a high goodness-of-fit (R²=0.959), as shown in Figure 4C. This particular form of degree distribution is indicative of the presence of highly connected regions, commonly referred to as hubs, within the GWM-HFN. To identify central regions, we defined hubs as the top 15% of nodes with the highest degree^31,32^. This data-driven approach identified 14 hub nodes. The specific anatomical locations and network affiliations of these 14 hubs are detailed in Table S4 and visualized in Figure 4D. These hubs are predominantly located in higher-order association cortices and primary sensory areas, including key regions within the default mode, visual, and limbic networks.

### 2.5. Comparison with Traditional GM-GM Networks (SLIM Dataset)

#### 2.5.1. Similarity Assessment

To evaluate the added value of incorporating white matter in connectivity modeling, we compared the GWM-HFN with traditional GM-GM networks at the edge, network, and global levels using the SLIM dataset. At the edge level, the two network types exhibited strong similarity, with an average Pearson correlation coefficient of 0.77 (±0.038). As shown in Figure 5A, a representative participant’s data revealed a high correlation between GWM-HFN and GM-GM connection strengths (r = 0.743). Nevertheless, this suggests that more than 40% of the variance remains unique to each network type. To better understand these differences, we identified the 5% of edges with the highest and lowest correlations between the two networks, illustrated in Figures 5B and 5C. Edges with low correlations were primarily intra-network connections within specific functional systems such as the VN, FPN, and DMN. In contrast, highly correlated edges were predominantly inter-network connections, especially between the BGN and other systems like the VN, SMN, LB, and DMN. This trend was consistent at the network level: inter-network correlations across large-scale functional networks were stronger (mean r = 0.84) than intra-network correlations (mean r = 0.79), as depicted in Figure 5D. At the global level, GWM-HFN and GM-GM connectivity profiles showed robust consistency across participants, with a global correlation coefficient of 0.857. Thus, these findings collectively reveal a critical pattern: while the two methods agree on broad, inter-system communication, the unique value of GWM-HFN lies in uncovering a more divergent topological structure within functional systems, providing novel insights into white matter-mediated integration.

**Figure 5.**
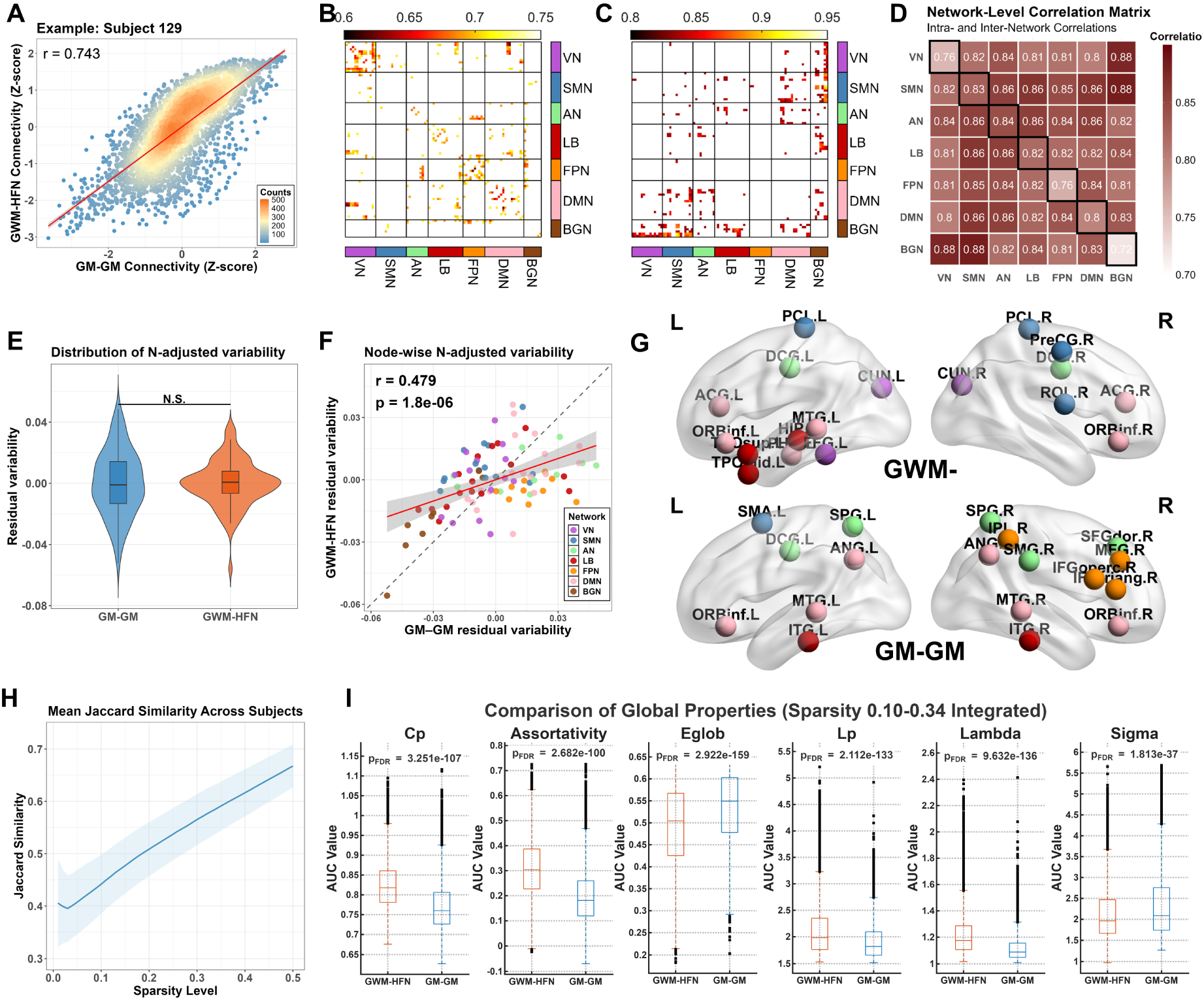
Comparative Analysis of GWM-HFN and GM-GM Networks. (A) Scatterplot illustrating the relationship between GWM-HFN and GM-GM connection strengths for a representative participant, showing a high correlation (r = 0.743). (B, C) Heatmaps highlighting edges with the lowest (<5th percentile) and highest (>95th percentile) correlations between the two network types, respectively. (D) Network-level correlation analysis revealing stronger inter-network correlations (mean r = 0.84) than intra-network correlations (mean r = 0.79) between GWM-HFN and GM-GM connectivity. (E) Violin plots comparing the magnitude of "true" inter-subject variability between the two frameworks after correcting for intra-subject measurement noise (N). No significant difference was found (Wilcoxon signed-rank test, p = 0.73). (F) Scatter plot illustrating the moderate correlation of node-wise "true" inter-subject variability between the GWM-HFN (y-axis) and GM-GM (x-axis) frameworks across 90 brain regions. The dashed line represents identity (y=x), and the solid red line is the linear regression fit. (G) Lateral views of the top 20% (18 out of 90) nodes with the highest "true" inter-subject variability for the GWM-HFN framework (top row) and the GM-GM framework (bottom row). The minimal overlap (Jaccard index = 0.125) highlights the distinct spatial topographies of variability captured by each method. (I) Line plot of average Jaccard coefficients across sparsity thresholds (0.01–0.50), with shaded regions representing standard deviation, demonstrating consistency across thresholds. (J) Box plots comparing the area under the curve (AUC) for global topological properties—including clustering coefficient, assortativity, characteristic path length, global efficiency, and small-worldness—across the 0.10–0.34 sparsity range, indicating significant differences between GWM-HFN and GM-GM networks.

#### 2.5.2. Variability Assessment

To compare the inter-individual variability captured by each framework while accounting for potential differences in signal quality, we applied variance decomposition method proposed by the Mueller et al. using our test-retest SLIM dataset^33^. This approach models the observed between-subject variability (**Y**) as a function of the intra-subject (session-to-session) variability (**N**), thereby allowing us to derive a residual measure of inter-subject variability that has been statistically corrected for measurement noise.

The analysis revealed that while the raw between-subject variability was initially higher in the GWM-HFN framework (mean Y = 0.711) than in the GM-GM framework (mean Y = 0.454), this was largely explained by a correspondingly higher level of intra-subject variability (i.e., noise; mean N = 0.631 vs. 0.368). After rigorously correcting for this intra-subject variability, the remaining "true" inter-subject variability was not significantly different in magnitude between the two frameworks (Wilcoxon signed-rank test, p = 0.73, Figure 5E).

Despite this comparable overall magnitude, the spatial topographies of this true variability were largely distinct. The node-wise variability maps were only moderately correlated (r = 0.479, 95% CI [0.302, 0.624], p = 1.80×10⁻⁶, Figure 5F), and the overlap between the top 20% most variable nodes was minimal (4 of 18 nodes; Jaccard index = 0.125, Figure 5G). Neuroanatomically, GWM-HFN’s high-variability nodes were concentrated in the DMN and SMN networks. In contrast, the GM-GM framework’s variability was highest in higher-order association systems, including the AN and FPN.

#### 2.5.3. Network Organization Assessment

To evaluate the organizational similarity between GWM-HFN and GM-GM networks, we computed Jaccard coefficients across a range of sparsity thresholds (0.01–0.50). The average coefficients ranged from 0.39 to 0.66, with values increasing at higher sparsity levels (Figure 5H), indicating greater structural alignment at higher network densities. At lower sparsity thresholds, notable discrepancies emerged, reflecting amplified structural differences between the two network types. This pattern was further nuanced across functional modules (Figure S1). Specifically, inter-network connections demonstrated lower baseline similarity, which progressively increased with sparsity, while intra-network connections showed higher baseline similarity but followed a U-shaped trajectory, reaching a minimum around 0.05 sparsity. These results suggest that WM-mediated interactions introduce distinct network architectural features, particularly in sparse functional networks, emphasizing the complementary contributions of GWM-HFN connectivity to traditional gray matter-based analyses.

To more comprehensively assess topological differences, we performed a graph-theoretical analysis using area under the curve (AUC) metrics calculated from the 0.10–0.34 sparsity range, with statistical comparisons based on paired t-tests. Results revealed that GWM-HFN networks are topologically distinct, exhibiting enhanced modular segregation but reduced global integration compared to GM-GM networks. GWM-HFN networks showed significantly higher Clustering Coefficient (Cp) and Assortativity, reflecting increased local cohesiveness and node-type homogeneity (Figure 5I). Specifically, after correcting for multiple comparisons, Nodal Clustering Coefficient and Subgraph Centrality in the GWM-HFN framework were significantly higher across a majority of brain areas. We found that 60 of 90 nodes for Clustering Coefficient and 76 of 90 nodes for Subgraph Centrality showed significantly greater values (paired t-test, P_FDR_ < 0.001), with detailed node-wise results presented in Figure S2. Lower Participant Coefficient in GWM-HFN networks suggested stronger modular boundaries, implying that white matter-informed connectivity preserves distinct community structures more rigidly (Figure S2). In contrast, GWM-HFN networks demonstrated significantly longer Characteristic Path Lengths (Lp) and lower Global Efficiency (Eglob), consistent with reduced capacity for rapid and efficient global information transfer (Figure 5J). This was associated with reduced Small-Worldness (Sigma), reflecting a diminished balance between segregation and integration. While Betweenness Centrality was significantly elevated in many GWM-HFN nodes, indicating increased control over information flow, other metrics—including Degree, Eigenvector Centrality, PageRank Centrality, and Nodal Efficiency—exhibited bidirectional changes across brain regions, suggesting a complex reorganization of functional hubs and peripheral nodes (Figure S2). Collectively, these findings demonstrate that GWM-HFN networks encode a distinct functional topology, marked by heightened modularity and segregation but less efficient global integration, suggesting a functional architecture that prioritizes specialized, modular processing over efficient global integration.

To directly compare the central network architectures, we applied the identical hub identification procedure (top 15% degree) to the conventional GM-GM networks (Figure 4E). As detailed in Table S4, this comparison revealed a substantial overlap, with 11 hubs being common to both methods (Dice coefficient = 0.786). These common hubs, including key regions in the bilateral middle temporal gyrus and lingual gyrus, form a robust core set of highly connected nodes identifiable by both approaches. Critically, the analysis also revealed method-specific hubs. The GWM-HFN uniquely identified three hubs in higher-order association areas: the right inferior frontal orbital gyrus (ORBinf.R), the left superior temporal gyrus (STG.L), and the right superior temporal pole (TPOsup.R). Conversely, the conventional GM-GM method uniquely identified three hubs located primarily in primary sensory and attention networks: the right superior frontal gyrus (SFGdor.R), the left calcarine cortex (CAL.L), and the right fusiform gyrus (FFG.R). In summary, this direct comparison demonstrates that while both frameworks identify a core set of shared hubs, they also reveal distinct topological priorities, with GWM-HFN highlighting unique hubs in higher-order association cortices and the conventional method in sensory and attention networks.

### 2.6. Age-Related Patterns in GWM-HFN (SALD datasets)

Using the SALD datasets, which include a broad adult age range from 19 to 80 years, we examined how GWM-HFN connectivity evolves across the adult lifespan. To comprehensively evaluate the unique characteristics of GWM-HFN, we benchmarked our findings against a parallel analysis of traditional GM-GM functional connectivity. Linear and quadratic regression models were applied to assess the relationship between age and connectivity strength, with head motion and sex controlled as covariates. At the global level, GWM-HFN connectivity showed a significant negative linear association with age (r = -0.252, p < 0.001; Figure 6A), while the quadratic term was not statistically significant (p = 0.296), indicating a predominantly linear age-related decline. In contrast, the benchmark GM-GM analysis revealed a significant inverted U-shaped quadratic relationship with age at the global level (β^qud^ = -0.017, p = 0.030; Figure 6D).

**Figure 6.**
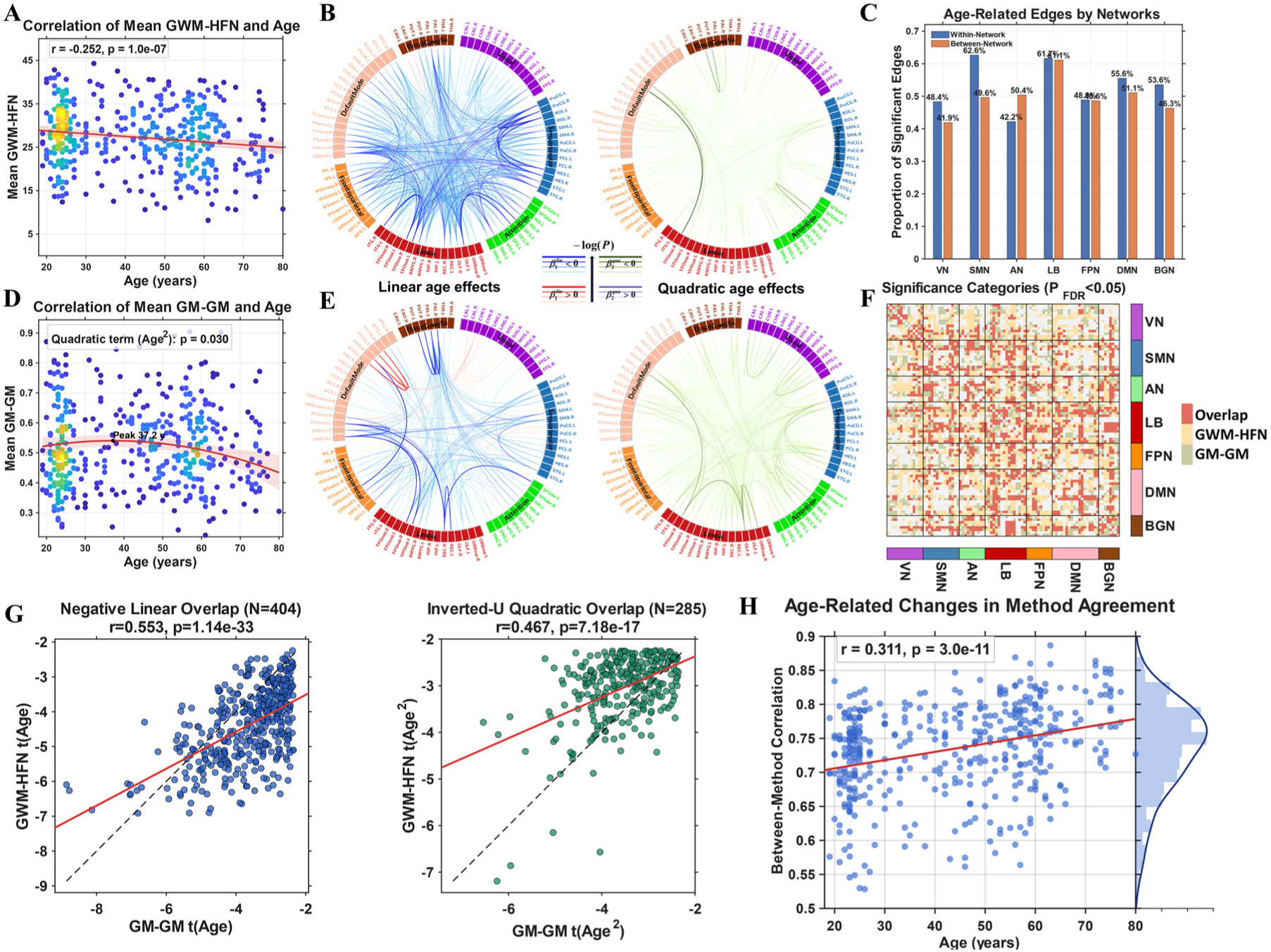
Age-Related Patterns of GWM-HFN and GM-GM Connectivity and Their Direct Comparison. (A) and (D) Scatter plot illustrating the negative linear correlation between mean GWM-HFN connectivity and age (r = -0.252, p = 1.0 × 10⁻^07^), while (D) shows the contrasting inverted U-shaped quadratic relationship for the benchmark GM-GM network. In both plots, each dot represents an individual, and its color indicates the density of overlapping data points. The red line depicts the fitted linear regression. (B) and (E) Circular connectograms depicting edge-level age effects for GWM-HFN and GM-GM networks, respectively. For each, the left panel shows linear effects (blue: negative, red: positive), and the right panel shows quadratic effects (green: inverted U-shaped, purple: U-shaped), with line thickness representing effect magnitude. Brain regions are color-coded according to their associated functional networks. (C) Clustered bar chart showing the proportion of significant age-related edges for the GWM-HFN framework, distributed across the seven functional networks and separated into within-network and between-network connections. (F) Heatmap illustrating the overlap of significant age-sensitive edges between the two frameworks. Red indicates edges significant in both methods (overlap), yellow indicates edges specific to GWM-HFN, and green indicates edges specific to GM-GM. (G) Scatterplots showing the significant positive correlation between the effect sizes (t-values) of overlapping age-sensitive edges for negative linear effects (left panel) and inverted U-shaped quadratic effects (right panel). (H) Scatter plot showing the positive correlation between age and the similarity of GWM-HFN and GM-GM connectivity patterns (r = 0.311, p = 3.0× 10⁻^11^), indicating greater convergence between the two methods in older individuals.

At the edge level, our GWM-HFN framework identified that over half of all connections (2044 out of 4005; 51.04%) were significantly modulated by age after FDR correction (*P*_*FDR*_ < 0.05). Notably, 78.47% of these age-sensitive edges (1604 connections) exhibited negative linear associations, suggesting widespread weakening of GWM-HFN connectivity with aging. A smaller proportion (1.96%, 40 edges) showed positive linear trends, indicating a limited set of connections that strengthen over time (Figure 6B, left panel). In addition to linear effects, quadratic age-related patterns were identified in 400 edges (19.57%). Of these, 96% (384 edges) followed inverted U-shaped trajectories, peaking before declining, whereas 4% (16 edges) followed U-shaped curves with initial decline followed by later-life increases (Figure 6B, right panel). The average peak age for connections with significant quadratic effects was 33.84 years, suggesting a critical window of maximal GWM-HFN in early adulthood. At the large-scale functional network level, we further investigated whether age-related effects preferentially targeted connections within or between functional networks by comparing the proportions of affected edges in each category. Overall, we observed that a substantial proportion of both within-network (55.4%) and between-network (51.5%) connections were significantly modulated by age, indicating a widespread impact. However, the analysis revealed network-specific patterns. For instance, the SMN and LB showed a higher proportion of affected within-network connections (62.6% and 61.7%, respectively) compared to their between-network counterparts (49.6% and 61.1%). Conversely, the AN exhibited a greater proportion of affected between-network connections (50.4%) relative to its within-network connections (42.2%). These findings, detailed in Figure 6C, suggest a heterogeneous pattern of age-related changes across the connectome rather than a uniform targeting of between-network links.

The benchmark GM-GM analysis, in turn, identified 1550 connections with significant age-related effects (*P*_*FDR*_ < 0.05), as illustrated in Figure 6E. Of these, 656 edges showed a linear relationship (499 negative, 157 positive), while a larger proportion (894 edges) followed a quadratic trajectory (889 inverted U-shaped, 5 U-shaped), with an average peak age of 38.44 years. A direct comparison of the age-sensitive edges identified by both methods revealed a substantial overlap of 938 connections (Jaccard index = 0.3532; Figure 6F). Within this overlap, consistency was high: 412 edges were linear (with 411, or 99.8%, showing the same direction of association) and 287 were quadratic (all sharing the same shape). Furthermore, the effect sizes of these common connections were significantly correlated between the two frameworks for both negative linear (r = 0.553, p < 0.001) and inverted U-shaped effects (r = 0.467, p < 0.001; Figure 5G). When categorizing effects more broadly into declining (negative linear, inverted-U) versus inclining (positive linear, U-shape) patterns, we found that 921 of the 938 overlapping edges (98.2%) showed this general consistency, indicating that when both frameworks detect an age-related change, they are often capturing a shared underlying biological process.

Finally, beyond the specific overlap in age-sensitive edges, we examined the overall similarity between the two connectome types across all subjects. We observed a high correlation between GWM-HFN and traditional GM-GM connectivity across subjects (mean r = 0.735), which intensified with increasing age (Figure 6H). This finding suggests that the convergence between GWM-HFN and gray matter-based functional networks becomes more pronounced in older individuals.

### 2.7. Clinical Relevance: Insights from Patient Cohorts

To test the clinical utility of the GWM-HFN framework, ASD was selected as the primary clinical exemplar for this study. This choice was motivated by several key theoretical and empirical considerations. First, ASD has long been conceptualized as a disorder of atypical brain connectivity, or a ‘disconnection syndrome,’ with numerous studies reporting altered functional connectivity patterns during tasks related to complex cognitive and social processing^34^. Second, there is extensive and well-documented evidence from neuroimaging studies demonstrating widespread alterations in white matter microstructure in individuals with ASD^35,36^. These established structural abnormalities provide a strong a priori basis for hypothesizing that WM-mediated functional connectivity, as captured by GWM-HFN, would also be affected. Finally, the publicly available ABIDE-II project offers a large-scale, multi-site dataset with rich clinical phenotyping, providing the necessary statistical power for a robust validation of our method and for exploring crucial brain-behavior relationships. To assess these potential alterations, we constructed networks for individuals with ASD and TC individuals using the ABIDE-II dataset, a large-scale, multi-site repository that integrates neuroimaging data from diverse international cohorts.

Prior to analysis, we applied the ComBat method for harmonization to minimize the impact of sites^37^. While a visual inspection of the distributions before and after harmonization suggests a reduction in site-related variance (Figure 7A), we performed a rigorous quantitative analysis to confirm the complete removal of site effects. Before harmonization, a Type III ANOVA revealed a highly significant site effect on global mean connectivity (F(12, 623) = 9.65, p < 2.2 × 10^-16^). At the edge-level, 93.8% of all connections showed a significant site effect (*P* < 0.05), with a median effect size (partial η^2^) of 0.086. After ComBat harmonization, the site effect was no longer significant at the global level (F(12, 623) = 0.67, p = 0.783). Critically, at the edge level, the proportion of connections with a significant site effect dropped to 0, and the median partial η^2^ was reduced to 0.002. These quantitative results confirm that the ComBat procedure effectively and comprehensively removed site-specific biases from our data.

**Figure 7.**
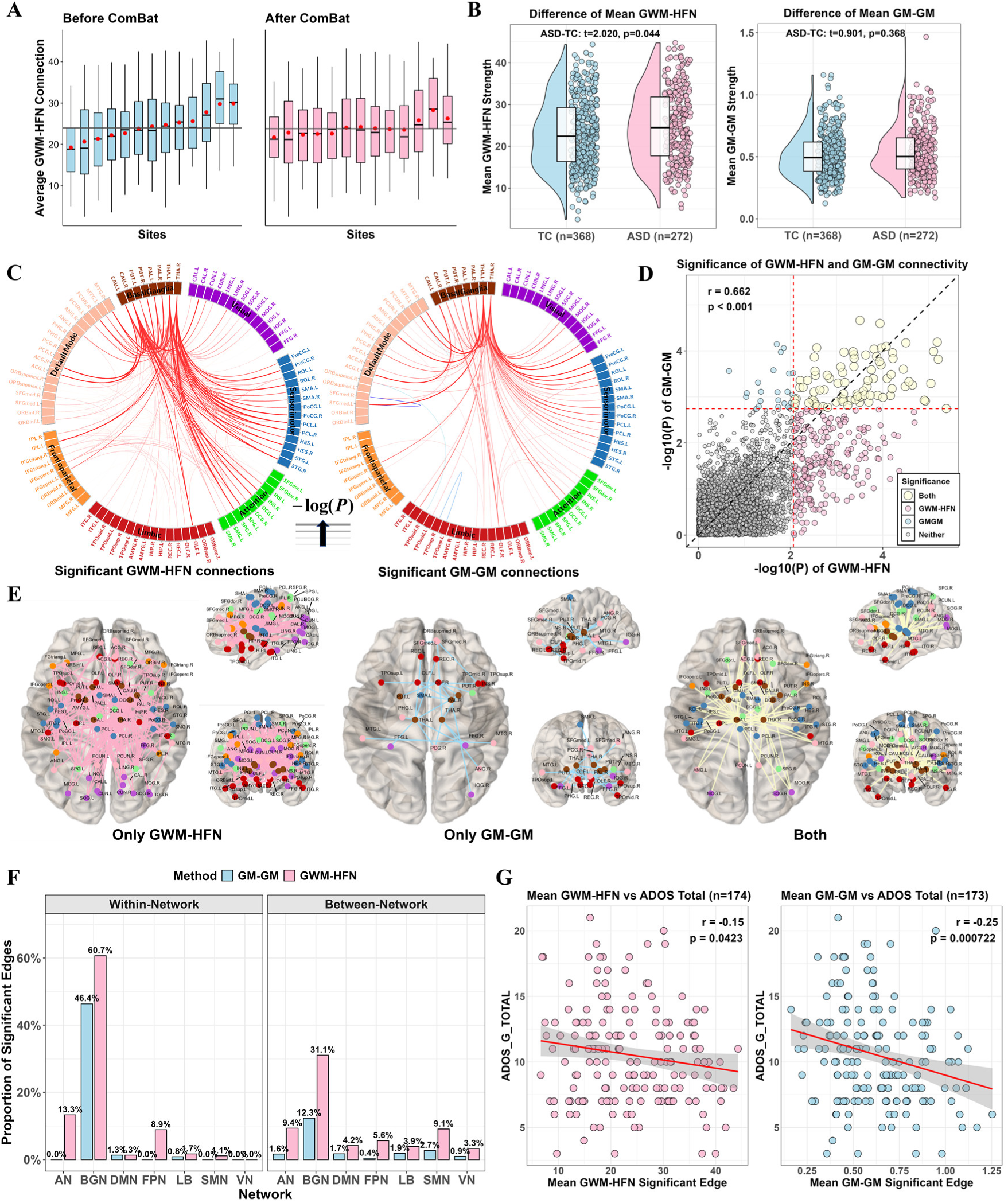
Comparative Analysis of GWM-HFN Connectivity Between ASD and TC Groups. (A) Boxplot illustrating the global mean GWM-HFN connectivity across 13 acquisition sites before and after harmonization. The red diamond indicates the mean connectivity within each site, while the gray line represents the overall mean connectivity across all sites. (B) Raincloud plot comparing the global-averaged GWM-HFN and GM-GM connectivity strengths between the ASD and TC groups. (C) Left panel: Brain network depicting the significantly different GWM-HFN connections identified between the ASD and TC groups, with all significant connections showing greater connectivity in the ASD group (ASD > TC), indicated by red. Right panel: Brain network illustrating the significantly different GM-GM connections identified between the ASD and TC groups, with the majority of connections exhibiting greater connectivity in the ASD group (ASD > TC), indicated by red. Connections showing decreased connectivity in the ASD group (ASD < TC) are indicated by blue. Line thickness corresponds to the magnitude of the effect size. (D) Scatterplot comparing the p-values of group differences for GWM-HFN and GM-GM functional connections. The two red dashed lines indicate the threshold for significance after FDR correction (*P*_*FDR*_ < 0.05). Data points are color-coded based on the significance of the connection: light yellow for connections significant in both the GWM-HFN and GM-GM analyses, light red for connections significant only in the GWM-HFN analysis, and light blue for connections significant only in the GM-GM analysis. (E) Brain connectivity map visualizing connections that were significant only in the GWM-HFN analysis, significant only in the GM-GM analysis, and significant in both analyses. (F) Bar plots quantifying the distribution of significant edges from the ASD versus TC group comparison, comparing the number of altered within-network and between-network connections for both the GWM-HFN and GM-GM frameworks. (G) Scatterplot showing the significant negative Pearson correlation between the mean connectivity strength of the altered edges and the ADOS-G total scores for both the GWM-HFN and GM-GM frameworks.

We first conducted group comparisons, controlling for age, sex, and mean FD as covariates. Results revealed that individuals with ASD exhibited significantly stronger global mean GWM-HFN connectivity compared to TC (*t* = 2.055, *p* = 0.042). In contrast, no significant differences were observed in mean GM-GM connectivity between the groups, as illustrated in **Figure 7B**. At the edge level, 310 GWM-HFN connections showed significant group differences, all indicating increased connectivity in the ASD group (**Figure 7C**). In comparison, the GM-GM connectivity network revealed only 106 significant edges, of which 102 also showed stronger connectivity in ASD. Remarkably, about 80% (83 out of 106) of these significant GM-GM connections overlapped with the GWM-HFN connections (**Figures 7D & 7E**), underscoring the heightened sensitivity of GWM-HFN in detecting ASD-related alterations.

Subsequently, we compared the proportions of significant connections to provide an unbiased assessment of within-versus between-network effects. This analysis revealed that ASD-related abnormalities were not predominantly between-network, but were instead highly concentrated within specific functional systems. This pattern was most pronounced in the BGN, where the proportion of affected intra-network connections was dramatically higher than between-network connections for both GWM-HFN (60.7% vs. 31.1%) and GM-GM networks (46.4% vs. 12.3%). These findings indicate that ASD-related hyperconnectivity is primarily characterized by a disruption of intra-network integrity, particularly within the basal ganglia—an effect to which the GWM-HFN framework showed heightened sensitivity (Figure 7F).

Finally, we investigated whether the identified significant connections were associated with the severity of autistic symptoms, as measured by the Autism Diagnostic Observation Schedule-Generic (ADOS-G). Specifically, we calculated the correlation between the total ADOS-G score (ADOS_G_TOTAL) and the mean connectivity strength of the identified GWM-HFN and GM-GM connections. This association was significant for the GWM-HFN framework (r = -0.154, p = 0.0423) and was notably more robust for the conventional GM-GM network (r = -0.255, p = 0.000722), as illustrated in **Figure 7G**.

### 2.8. GWM-HFN Correlations with Cognitive and Behavioral Measures

The BGSP dataset included a broad array of behavioral and cognitive measures, enabling exploration of phenotypic correlates of GWM-HFN connectivity. To investigate these associations, we employed partial least squares (PLS) regression to evaluate whether GWM-HFN connectivity could explain interindividual differences in four cognitive-behavioral phenotypes: Shipley vocabulary score, matrix reasoning scores, and estimated IQ derived from both Shipley-Hartford T-scores and the OPIE3 formula. A parallel PLS analysis was conducted using the conventional GM-GM networks as a direct baseline comparison.

Our analysis focused on the first PLS component (PLS1), which represents the linear combination of GWM-HFN connectivity features most strongly associated with the behavioral data. Using permutation testing with 10,000 permutations, we found that PLS1 scores explained a significant proportion of variance in both the Shipley vocabulary task (8.95%, *P_permutation_* = 0.0229, **Figure 8A**) and the Shipley-Hartford estimated IQ (9.09%, *P_permutation_* = 0.0186, **Figure 8B**). For the benchmark GM-GM analysis, PLS1 scores showed a significant association only with the Shipley vocabulary task (13.05 %, *P_permutation_* = 0.0060, **Figure 8C**). No significant associations were found for the matrix reasoning score or IQ estimated using the OPIE3 formula for either framework. Notably, the GWM-HFN was unique in identifying a significant link to the Shipley-Hartford estimated IQ, an association that was not detected by the conventional GM-only analysis.

**Figure 8.**
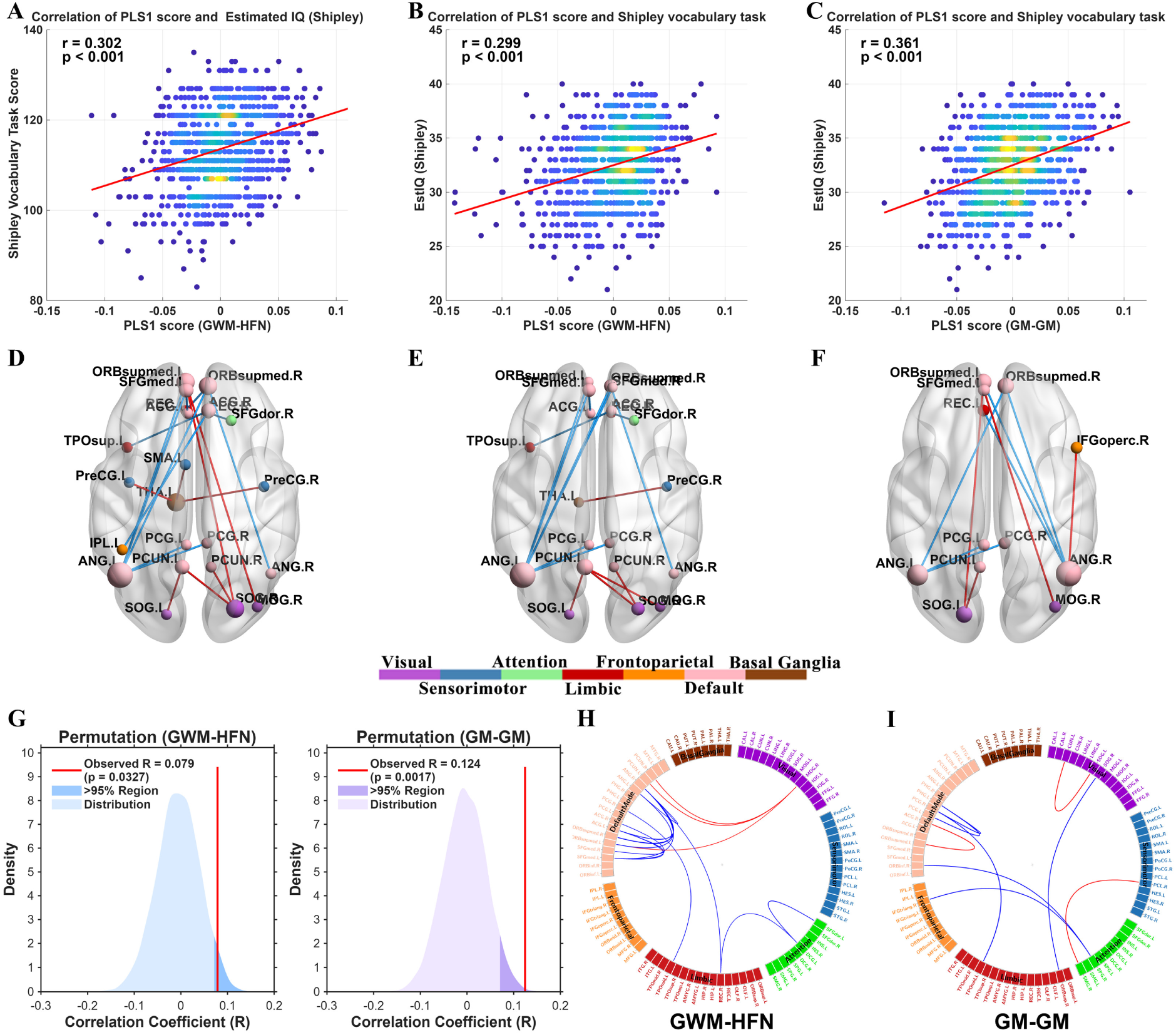
Association between GWM-HFN connectivity and Behavior and Cognition measures. (A, B, C) Scatterplots illustrating the significant correlations between PLS1 scores and cognitive measures. (A) Correlation between GWM-HFN PLS1 scores and the estimated IQ derived from Shipley-Hartford Age-Corrected T-Scores. (B) Correlation between GWM-HFN PLS1 scores and the Shipley vocabulary task scores. (C) Correlation between the benchmark GM-GM PLS1 scores and the Shipley vocabulary task scores. The color of the data points in the scatterplots indicates the density of overlapping points, with warmer colors representing higher density. (D, E, F) Brain visualizations of the stable, high-weight connectivity patterns driving the significant PLS associations shown in A, B, and C, respectively. Edges displayed are those with absolute standardized PLS1 weights exceeding 3σ and confirmed to be stable via bootstrap analysis. (G) Density plots of 10,000 permutation tests for the correlation coefficient (r) between connectome data and Shipley vocabulary task scores using the BBS modeling method. The results demonstrate significant individual-level prediction for both the GWM-HFN (left panel) and GM-GM (right panel) frameworks. (H, I) Circos plots visualizing the key connectivity patterns with the highest contributions (|z| > 3σ) to the significant BBS predictions for the GWM-HFN (H) and GM-GM (I) vocabulary models. Positive weights are shown in red, and negative weights are in blue.

To identify the most robust connectivity patterns driving these effects, we examined the standardized PLS1 weights and assessed the stability of all edge weights using a bootstrap analysis. For subsequent interpretation and visualization, we focused on high-weight connections (absolute standardized weight (|z| > 3σ) that were also confirmed to be highly stable (Bootstrap Ratio |BSR| > 2). For the GWM-HFN framework, the stable, high-weight connectivity patterns underlying both the Shipley vocabulary and the Shipley-Hartford estimated IQ associations were remarkably consistent (Figure 8D&E). They were characterized by predominantly negative weights within the DMN and positive weights between the DMN and the VN, with a high overlap between the contributing edges for both measures (Jaccard index = 0.62). Intriguingly, the benchmark GM-GM network’s significant association with vocabulary was driven by the same neurobiological pattern—negative intra-DMN and positive DMN-VN connectivity (Figure 8F). A direct comparison revealed that half (5/10) of the high-weight edges from this GM-GM analysis were also identified by the GWM-HFN for the vocabulary task, indicating a shared substrate captured by both methods.

To further validate these associations, we applied the brain bias set (BBS) modeling method^38^ to predict individual scores in the cognitive domains identified as significant in the PLS analysis. Briefly, this approach uses principal component analysis to reduce the dimensionality of the connectivity data before fitting a linear regression model within a cross-validation framework. Results showed that GWM-HFN connectivity significantly predicted individual scores in the Shipley vocabulary task (mean *r* = 0.0785, *P_permutation_* = 0.0327, Figure 8G), while no significant prediction was found for IQ estimated using the Shipley-Hartford T-scores. The benchmark GM-GM network also significantly predicted vocabulary scores (mean *r* = 0.124 *P_permutation_* = 0.0017). Finally, we visualized the connectivity patterns with the highest contributions (|z| > 3σ) to these significant predictions for the GWM-HFN and GM-GM frameworks, as depicted in Figure 8H&I, respectively.

## 3. Discussion

This study introduces and rigorously validates the GWM-HFN, a novel framework that reliably integrates WM BOLD signals into functional connectome analysis. Our multi-dataset validation demonstrates GWM-HFN’s robust test-retest reliability, distinct topological features, and relevance in capturing age-related and clinical alterations. Such comprehensive characterization across reliability, topology, and diverse applications addresses a critical need in the evolving field of unified GM-WM functional connectomics, where extensive validation of emerging models^24,25^ is still developing. By providing a validated means to incorporate WM’s functional contributions, GWM-HFN directly challenges the traditional GM-centric view in neuroimaging^2,7^. Specifically, its GM-centered representation, derived from GM-WM covariance, overcomes limitations of WM-only approaches^13,17^ and bipartite GM-WM models that hinder conventional global network analyses due to absent closed-loop interactions ^24^. GWM-HFN therefore offers a more holistic and interpretable model of whole-brain functional architecture, significantly advancing our capacity to map complex neural communication.

### 3.1 Reliability of GWM-HFN in the Context of Resting-State fMRI

A cornerstone of establishing the GWM-HFN framework’s utility lies in its methodological viability, particularly its test-retest reliability. Our analyses yielded mean edge-wise ICCs of approximately 0.36 for short-term (BNU-3 dataset) and 0.20 for long-term (SLIM dataset) GWM-HFN connectivity. According to benchmarks such as those proposed by Landis and Koch^39^, these ICCs position GWM-HFN reliability in the ‘fair’ (ICC = 0.21–0.40) and ‘slight’ to ‘fair’ (ICC ≈ 0.20) categories, respectively. These values must be interpreted within the challenging context of rs-fMRI research, where achieving high test-retest stability for any individual FC measure is non-trivial due to inherent physiological noise, ongoing unconstrained cognitive processes, and cross-study analytical variability^28,29^.

Our comprehensive validation now provides crucial context for these values through a direct comparison with conventional GM-GM networks. We found that the ICCs for GWM-HFN, while within the spectrum reported for many GM-based connectivities^28^, are significantly lower than those of the GM-GM network. To understand the implications of this finding, we adopted a more comprehensive assessment framework^30^ which revealed a critical trade-off between raw stability and individual specificity. While the GM-GM network demonstrated higher raw stability (i.e., higher ICC and Reliability, µ_intra_), our GWM-HFN framework showed a marked advantage in its ability to capture individual uniqueness. This was evidenced by its significantly lower Uniformity (µ_inter_), indicating that GWM-HFN connectomes are more distinct from one another across the population.

We propose that the superior stability of GM-GM networks is largely driven by strong, generic signals from primary sensory-motor regions that are highly consistent both within and across individuals. In contrast, GWM-HFN, by design, leverages the covariance of interaction profiles with all WM bundles, a method that is more sensitive to the subtle and variable—yet more individually-specific—connectivity patterns of higher-order association networks. The assessment of reliability in methodologies incorporating WM signals is an area of active development. For instance, Wang et al. investigated the robustness and reliability of direct WM functional connectivity^40^, finding that while static FC in GM networks was stronger than in WM networks, WM networks exhibited greater dynamism. The GWM-HFN approach, by deriving connectivity from shared WM bundles utilization, possesses distinct reliability characteristics. Our findings suggest that its strength lies not in maximizing raw signal stability, but in its enhanced sensitivity to the trait-like, neurobiological variations that differentiate individuals.

Furthermore, the pursuit of enhanced reliability underscores the critical importance of standardized preprocessing pipelines, as highlighted by the development of specialized toolboxes like WhiFuN^11^. Therefore, the reliability profile demonstrated by GWM-HFN is a crucial validation. It establishes that our framework, while exhibiting lower raw stability than GM-GM networks, successfully captures a rich source of individual-specific variance. This superior ability to characterize individual differences provides a necessary foundation for employing GWM-HFN in longitudinal studies and for investigating its potential in biomarker development, where tracking unique individual features is paramount^10,14,15^.

### 3.2 Distinct Topological Organization of WM-Mediated Functional Networks

The GWM-HFN framework not only demonstrates reliability but also unveils a unique topological signature for WM-mediated functional networks. These networks inherently possess characteristics of efficient organization, such as small-worldness, modularity, and hub structures, as established by our graph-theoretical analyses. More critically, when contrasted with conventional GM-GM networks, GWM-HFNs exhibit a distinct profile: enhanced functional segregation alongside reduced global integration efficiency. This topological shift is a direct consequence of the GWM-HFN methodology, which defines GM-GM functional links based on their shared covariance profiles with WM tracts. Such an approach naturally groups GM regions that functionally ‘resonate’ with the same WM bundles, thereby strengthening intra-modular cohesion and revealing a functional architecture more explicitly sculpted by the brain’s WM scaffolding^41–43^. The resulting increase in modularity and local clustering suggests that GWM-HFN can delineate functional communities whose integrity is fundamentally tied to specific WM conduits.

The observed reduction in global efficiency and increased path length in GWM-HFNs, relative to GM-GM networks, likely signifies the multi-step nature of WM-mediated communication (GWM-HFN paths) versus potentially more direct GM-GM BOLD synchrony. This implies that while WM bundles facilitate specific and organized communication, the overall information transfer across the entire GWM-HFN might appear less direct or rapid when primarily channeled through these anatomically constrained routes^24,25^. This trade-off results in a modified small-world architecture, potentially favoring specialized processing within modules at the cost of global broadcasting efficiency.

Furthermore, the GWM-HFN framework reshapes the functional importance of nodes; our direct comparison revealed that while both frameworks identify a core set of shared hubs in regions like the middle temporal gyrus, GWM-HFN uniquely identified hubs in higher-order association areas (e.g., inferior frontal orbital gyrus) that were not prominent in the GM-GM network. Conversely, the conventional method highlighted hubs primarily in sensory and attention networks. This suggests that the elevated betweenness centrality of certain GWM-HFN hubs makes them critical control points for inter-modular communication specifically within this WM-mediated system, distinct from hubs identified in GM-GM networks that may simply reflect high overall synchrony. Thus, the topological characteristics of GWM-HFN provide a more mechanistic view of functional organization, emphasizing how WM bundles structure and constrain neural communication.

### 3.3 Complementary Insights Beyond Conventional GM-GM Connectivity

While the GWM-HFN framework introduces a novel perspective by incorporating WM signals, its relationship with traditional GM-GM functional connectivity provides crucial context. Our findings indicate a considerable overlap (mean edge-level r≈0.77, SLIM dataset), affirming that GWM-HFN captures fundamental patterns of synchronous neural activity. However, the more than 40% of unique variance captured by GWM-HFN underscores its capacity to reveal aspects of functional organization not apparent in GM-only analyses. This is particularly evident in the divergent similarity patterns between the two frameworks across network sparsity thresholds. The U-shaped similarity for intra-network connections suggests that while both methods capture the strongest ‘core’ links, they diverge on moderately-strong connections. In contrast, the progressive increase in inter-network similarity suggests that the frameworks converge on capturing broader patterns of inter-system communication after initially differing on the most specific long-range links. This highlights GWM-HFN’s unique sensitivity to specific WM-mediated pathways, particularly at sparse network densities.

A key contribution of GWM-HFN lies in its ability to capture distinct patterns of inter-individual variability. Our initial analyses suggested heightened variability in GWM-HFN, particularly in connections involving the basal ganglia and limbic system. However, a more rigorous variance decomposition analysis, which corrected for session-to-session measurement noise, revealed a more nuanced picture^33^. After accounting for noise, the overall magnitude of "true" inter-subject variability was not significantly different between the GWM-HFN and GM-GM frameworks. Critically, though, the spatial topographies of this true variability were largely distinct. GWM-HFN’s high-variability nodes were concentrated in the DMN and SMN, whereas the GM-GM framework’s variability was highest in higher-order association systems like the AN and FPN. This finding strongly suggests that the unique variance captured by GWM-HFN is not simply attributable to higher noise but rather taps into a genuine and spatially distinct source of biological differences related to individuals’ WM-mediated communication profiles^44–46^. Such sensitivity is paramount for fields like developmental neuroscience and clinical research, where understanding individual trajectories and susceptibility is key^47^.

These differences in captured variance and sensitivity to individual differences are also reflected in the distinct network topology of GWM-HFN compared to GM-GM networks, as detailed previously (enhanced modular segregation and reduced global integration efficiency). Such topological distinctions are a natural consequence of a network model that explicitly accounts for WM-mediated interactions, aligning with emerging perspectives that WM signals actively contribute to the organization and potential ‘rewiring’ of GM functional communities^26^. Thus, GWM-HFN provides not just a similar picture to GM-GM networks but a complementary one, enriched by the functional information carried by WM functional connectivity profiles and offering a more nuanced understanding of brain-wide communication.

### 3.4 Age-Related Decline and Reorganization of GWM-HFN Connectivity

Applying the GWM-HFN framework across the adult lifespan (SALD dataset) revealed significant age-related dynamics, predominantly characterized by a linear decline in global mean connectivity strength. This overarching negative trajectory aligns well with the established literature documenting widespread age-related degradation in both structural WM integrity^48,49^ and conventional GM functional connectivity^5^. It also resonates with findings from related WM-GM network models, such as the reduced global efficiency observed with increasing age in bipartite WM-GM networks^24^. This contrasts with the benchmark GM-GM analysis, which revealed an inverted U-shaped global trajectory, suggesting that WM-mediated connectivity may follow a more direct path of age-related decline.

Beyond this global trend, GWM-HFN demonstrated sensitivity to more complex, edge-specific age trajectories. While the majority of affected connections exhibited linear decreases, reinforcing the theme of age-related weakening, a notable subset displayed inverted U-shaped quadratic patterns, despite the global quadratic term being non-significant. These connections typically peaked in early adulthood (mean peak age ∼34 years) before declining. The complexity of WM age-related changes is further highlighted by recent studies that reported both U-shaped patterns in static WM network connectivity strength and inverted U-shapes in the temporal variability of WM-GM connectivity across the adult lifespan^22^. This non-linear pattern is particularly compelling, as this peak age roughly coincides with the culmination of protracted maturation processes for certain cognitive functions and for WM microstructural properties, such as myelination^50^. The GWM-HFN may thus be capturing a critical window of maximal efficiency in WM-mediated functional communication during early to middle adulthood, followed by senescence potentially linked to the known microstructural deterioration of WM pathways later in life^49^. This finding supports the notion that certain localized functional systems undergo distinct developmental trajectories, a critical neurodevelopmental process that is obscured by global-level metrics.

Intriguingly, our findings (SALD dataset) also indicated that GWM-HFN and traditional GM-GM connectivity patterns become significantly more similar with advancing age^51^. This convergence could reflect several underlying processes. One possibility is age-related neural dedifferentiation, where diminished functional specialization of brain networks^52,53^ blurs the distinctions between connectivity reflecting direct GM synchrony and WM-mediated interactions, perhaps due to reduced specificity of WM-guided pathways or compensatory, more diffuse brain activity^54^. Alternatively, a ‘shared vulnerability’ scenario might explain this convergence, whereby common aging mechanisms like vascular or metabolic decline, and diffuse microstructural wear in both GM and WM^24,48,49^, could globally degrade both direct GM coupling and WM-pathway integrity, causing their functional signatures to appear more alike. Disentangling these potential contributors requires further research, but the GWM-HFN’s capacity to capture this age-related shift in network relationships, alongside broader linear declines and nuanced non-linear trajectories, highlights its value in comprehensively studying age-related functional brain changes and the evolving role of WM-mediated communication.

### 3.5 GWM-HFN Reveals Altered Connectivity in Autism Spectrum Disorder

This observed GWM-HFN functional hyperconnectivity likely relates to the atypical WM neurodevelopment and microstructure frequently reported in ASD^55,56^. While diffusion MRI studies often indicate reduced microstructural integrity (e.g., lower FA) across widespread WM tracts^57,58^, and morphometric analyses show heterogeneity but potential aggregation of volume reductions in common pathways^59^, the relationship between these structural characteristics and functional connectivity is intricate. The functional hyperconnectivity captured by GWM-HFN might signify compensatory mechanisms attempting to overcome less efficient signaling along structurally atypical pathways, or perhaps reflect altered excitatory/inhibitory balances specific to WM-mediated communication routes. The prominent involvement of BGN connections in our GWM-HFN findings aligns with reports of altered structural network topology involving the BGN even in preschool ASD^60^, hinting at early developmental atypicalities in cortico-striatal circuits relevant to ASD symptomatology^61,62^.

Crucially, the clinical relevance of these GWM-HFN alterations was substantiated by a significant negative correlation between the mean hyperconnectivity strength and total ADOS-G scores. While seemingly counterintuitive, this complex relationship (greater GWM-HFN hyperconnectivity associating with less severe symptoms) may reflect a successful compensatory mechanism rather than a primary pathological feature. Given the well-documented structural WM deficits in ASD^57,58^, this increased functional coupling could represent an adaptive neural response to overcome underlying inefficiencies. In this context, individuals with more severe symptoms may be those who fail to mount this compensatory response, a possibility consistent with the profound heterogeneity of the disorder^59^. Further research is needed to explore this connection across different symptom domains and developmental stages^58^.

While our current study focused on ASD, the established vulnerability of WM pathways across a range of psychiatric disorders^44^ suggests that sensitive functional probes like GWM-HFN could be valuable more broadly. For ASD specifically, GWM-HFN’s ability to detect robust functional alterations potentially missed by GM-GM analysis, coupled with its association with clinical severity, highlights its promise for complementing structural MRI and advancing our understanding of WM network dysfunction in this complex condition.

### 3.6 Linking GWM-HFN Connectivity to Individual Differences in Cognitive Performance

The functional significance of GWM-HFN was further underscored by its ability to explain interindividual variance in cognitive performance within the BGSP dataset. Initial PLS regression analyses revealed that GWM-HFN connectivity patterns were significantly associated with Shipley vocabulary scores and Shipley-Hartford estimated IQ. In a direct benchmark comparison, the conventional GM-GM network also showed a significant association with vocabulary but failed to detect a link to estimated IQ, highlighting the unique sensitivity of our WM-informed framework.

For the GWM-HFN framework, the stable, high-weight connectivity patterns underlying both associations were remarkably consistent. They were characterized by predominantly negative weights within the DMN and positive weights between the DMN and the VN, with a high overlap between the contributing edges for both measures (Jaccard index = 0.62)^63–65^. Intriguingly, the benchmark GM-GM network’s significant association with vocabulary was driven by the same neurobiological pattern— negative intra-DMN and positive DMN-VN connectivity. This indicates that both methods capture a shared, fundamental substrate for language-related cognition, while GWM-HFN provides additional sensitivity to broader cognitive functions like IQ.

To more rigorously test these brain-behavior relationships at an individual level, we employed the BBS modeling method, an approach designed to assess the predictive capacity of connectomic features for individual phenotypes^38^. Our application of BBS modeling provided crucial nuanced validation: GWM-HFN connectivity significantly predicted individual Shipley vocabulary scores, identifying specific edge contributions. This robust individual-level prediction for vocabulary, surviving cross-validation, strongly suggests that certain WM-mediated functional pathways captured by GWM-HFN are indeed tightly linked to this specific cognitive skill.

Interestingly, while the initial PLS analysis indicated an association with Shipley-Hartford estimated IQ, this did not translate into significant individual-level prediction under the BBS framework. This divergence is informative. PLS identifies broad patterns of covariance at the group level, whereas BBS provides a stricter test of individualized predictive utility. The lack of significant IQ prediction via BBS could suggest that either the true effect size for this particular IQ measure, while detectable by PLS, is insufficient for robust individual prediction with the current GWM-HFN feature set, or that the relationship between GWM-HFN connectivity and this IQ estimate is more complex and perhaps less directly linear than that observed for vocabulary. Sripada et al. themselves discuss how resting-state connectomes contribute to inter-individual variation^38^; it’s plausible that GWM-HFN features relevant to IQ are more diffusely represented or interact in a manner not fully captured by the specific BBS implementation used. Nevertheless, the successful individual prediction of vocabulary scores via GWM-HFN strongly indicates that this framework captures behaviorally relevant variance tied to specific WM-mediated communication pathways, highlighting its potential beyond group-level associations and underscoring that WM functional signals, as integrated by GWM-HFN, are indeed meaningfully related to cognitive function.

### 3.7 Limitations

Several limitations should be acknowledged when interpreting these findings. First, while we utilized longitudinal data for reliability assessment, the age-related and clinical analyses were primarily cross-sectional, precluding definitive inferences about developmental trajectories or causal relationships between network changes and clinical status. Future longitudinal studies employing GWM-HFN are warranted. Second, the BOLD signal in WM is inherently weaker and potentially more susceptible to physiological noise and partial volume effects than in GM. Although rigorous preprocessing steps, including WM-specific smoothing and nuisance regression strategies, were implemented, residual influences cannot be entirely excluded^11^. Continued methodological refinement for optimizing WM signal extraction and denoising remains important^10^. Third, our analysis focused on static functional connectivity. Exploring the dynamic fluctuations of GWM-HFN connectivity and its interplay with dynamic GM connectivity could offer further insights into how WM shapes transient neural coordination^22^. Finally, several methodological choices should be noted. Our use of anatomically-defined atlases (AAL, JHU-ICBM 48) represents a key methodological choice^68^. While this approach was deliberately chosen to ensure robustness and comparability across our several diverse datasets, we acknowledge that modern, functionally-defined parcellations, such as the Schaefer atlas^69^, may better align with the brain’s intrinsic functional organization. To address this, we performed a targeted validation analysis using the Schaefer-100 atlas, which confirmed that the fundamental topological properties of the GWM-HFN are robust to the choice of parcellation (see Figure S3). Nevertheless, a full replication of all our findings—including the lifespan, clinical, and cognitive analyses—using different parcellation schemes is an important avenue for future investigation. Furthermore, the GWM-HFN construction itself represents one specific approach to integrating heterogeneous signals. While our study provides a comprehensive benchmark against the traditional GM-only approach, a direct quantitative comparison with other emerging GM-WM network construction methods was not performed^24,25^. Establishing a fair comparison is complicated by the fundamentally different outputs of these alternative frameworks (e.g., directed graphs from bipartite projections, or three-dimensional tensors from three-way correlations) and remains a critical direction for future research. Similarly, we did not directly compare the GWM-HFN with structural connectivity metrics derived from diffusion MRI. Investigating the relationship between WM-mediated functional connectivity and the underlying anatomical connectome, to quantify the unique variance captured by each modality, remains a critical direction for future research.

### 3.8 Conclusion

In conclusion, this study introduces and comprehensively validates the GWM-HFN as a reliable and neurobiologically insightful approach that transcends the traditional GM-centric view by explicitly integrating WM functional signals. Our findings robustly demonstrate that GWM-HFN not only captures a distinct topological organization reflecting WM-mediated interactions ^41,43^ but also exhibits a superior ability to resolve the unique, individual-specific connectivity patterns that differentiate individuals, alongside significant relevance to lifespan changes, clinical conditions like ASD, and cognitive performance. By providing a validated novel lens on WM-mediated functional communication, the GWM-HFN framework offers a robust approach to better integrate the brain’s gray and white matter functional signals, thereby providing a more comprehensive model of whole-brain functional architecture. It thereby encourages a more holistic understanding of brain-wide communication, paving the way for refined investigations into the integral role of white matter in shaping the human connectome in both health and disease.

## 4. Methods and Materials

### 4.1. Datasets

To comprehensively evaluate the constructed GWM-HFN, we employed several independent datasets and conducted a systematic investigation from multiple perspectives. These analyses aimed to assess the networks’ test-retest (TRT) reliability, elucidate their organizational principles, compare them with traditional GM-GM networks, and examine their associations with aging trajectories, clinical relevance, and behavioral and cognitive correlates.

Specifically, the Southwest University Longitudinal Imaging Multimodal (SLIM) dataset (http://dx.doi.org/10.15387/fcp_indi.retro.slim) was employed to investigate long-term TRT reliability and network topology, while the Beijing Normal University (BNU-3) dataset (http://dx.doi.org/10.15387/fcp_indi.corr.bnu3) was used to assess short-term TRT reliability. The Southwest University Adult Lifespan (SALD) dataset (https://fcon_1000.projects.nitrc.org/indi/retro/sald.html) were utilized to examine age-related trajectories in GWM-HFN. The Autism Brain Imaging Data Exchange II (ABIDE-II) project (https://fcon_1000.projects.nitrc.org/indi/abide/abide_II.html) was analyzed to assess the clinical relevance of these networks. The Brain Genomics Superstruct Project (BGSP) dataset (https://www.neuroinfo.org/gsp/) was used to examine Behavioral and Cognitive Correlates of GWM-HFN. For a detailed description of these datasets, please refer to the Supporting Information (see “Study Datasets” section). **Table S3** provides a summary of the demographic details for each dataset.

### 4.2. Neuroimaging Preprocessing

Functional MRI images in all the datasets underwent the same analytical pipeline to construct GWM-HFN unless stated otherwise. All acquired images were subjected to preprocessing using a combination of the SPM12 toolbox (http://www.fil.ion.ucl.ac.uk/spm/software/spm12), DPARSF (http://rfmri.org/DPARSF) toolboxes.

Firstly, the individual T1 images were segmented into WM, GM, and cerebrospinal fluid (CSF) using SPM12’s New Segment algorithm and then normalized to the Montreal Neurological Institute (MNI) template. The rs-fMRI were removed the initial several time points, corrected for slice timing and head motion effects, and co-registered with the anatomical image. Subsequently, 24 motion-related parameters, the mean CSF signal and motion spikes were regressed out. Consistent with recommendations for studies investigating white matter function, global signal regression was not performed, as this procedure could artificially remove or distort the meaningful WM BOLD signals that are central to our analytical framework^10^. The data were detrended and temporally filtered with a passband frequency of 0.01 - 0.15 Hz. Then, spatial smoothing was performed separately in the WM and GM using a kernel with 4 mm full-width half-maximum (FWHM). Finally, the processed images were normalized to the standard MNI template and resampled to 3 mm³ voxels.

A manual quality control procedure was performed after preprocessing. The following criteria were used to assess data quality: (1) successful generation of all preprocessed results, (2) mean framewise displacement (FD) below 0.2 mm, and (3) visual inspection confirming satisfactory spatial normalization.

### 4.3. Construction of GWM-HFN Functional Connectome

As depicted in Figure 1, using the widely recognized AAL and JHU-ICBM 48 atlases, time series were extracted from 90 cortical GM regions (denoted as *g_i_*, *i* = 1, 2,…, 90) and 48 WM fiber bundles (denoted as w *_j_*, *j* = 1, 2, …, 48), derived from preprocessed functional imaging data. A 90×48 GM-WM functional correlation matrix was computed, representing the interaction between GM regions and WM tracts. In graph-theoretical terms, this GM-WM correlation matrix forms a bipartite graph 𝒢 = {*G*,*W*, *E*, *B*}, consisting of two distinct sets of nodes (*g _i_* ∈ *G*, *w _j_* ∈ *W*) where edges only connect nodes from different groups (with *E* representing the set of edges and *B* representing the edge weights). The lack of within-group connections (i.e., no WM-WM or GM-GM links) results in the absence of closed triangular paths, which complicates the evaluation of important network properties such as global efficiency^23^. To overcome this limitation, we propose a novel method that projects the original bipartite GM-WM network into a weighted unipartite network where only GM nodes are explicit, and WM nodes are implicit—referred to as the GWM-HFN.

Specifically, for each participant, the GM-WM correlation matrix (denoted as *B* = (*b_ij_*)_90×48_) is a 90 ×48 matrix where each row represents the functional connectivity of a GM region with multiple WM regions. To account for variability in connectivity strength and ensure comparability across different GM regions, we applied row-wise Z-score normalization to ***B,*** to normalize for overall connectivity strength differences across GM regions, yielding a normalized matrix ***Z***. This normalization is grounded in the biological principle that GM regions exhibit different levels of overall connectivity. By standardizing these values, the analysis emphasizes how each region interacts relative to its own connectivity profile, offering a clearer representation of the brain’s functional organization. Next, we computed the covariance matrix ***C*** of ***Z*′** to represent the functional connectivity between GM regions mediated by WM bundles. Each element of the resulting 90×90 matrix *C* = (*c_ij_*)_90×90_ = *Z* · *Z*′ quantifies the communication strength between cortical region **i** and cortical region **j** via shared WM connections. Thus, ***C*** serves as a GM FC matrix mediated by WM, also known as the GWM-HFN FC. This approach highlights the relative FC between GM regions, allowing for the detection of subtle interactions while accounting for variations in connectivity magnitude. It facilitates deeper insights into network properties and the investigation of clinically relevant connectivity changes.

### 4.4. Multi-level Test-Retest Reliability Analysis (BNU-3 and SLIM Datasets)

To evaluate the reliability of GWM-HFN FC, both short-term and long-term TRT reliability were assessed using the SLIM and BNU-3 datasets, respectively. To provide a comprehensive benchmark against conventional methods, all reliability analyses described below were performed in parallel for both the GWM-HFN and a standard GM-GM functional connectome.

First, we assessed the stability of the overall connectivity patterns. At the group level, we calculated the Pearson correlation between the group-averaged connectivity matrices from different time points. At the individual level, for each participant, we computed the Pearson correlation coefficients between their vectorized connectivity matrices from different time points and then averaged these coefficients across the group. This analysis addresses the discrepancy between group-level stability and individual-level consistency.

Furthermore, to quantify the reliability of individual connections, we utilized the intraclass correlation coefficient. A two-way random-effects model for single-measure agreement, ICC(2, 1), was employed, accounting for both individual variability and measurement consistency across time. Formally, for each edge in the GWM-HFN, the ICC was calculated as:

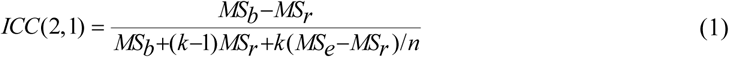

where MS_b_ is the mean square between subjects, MS_r_ is the mean square between measurements (time points), MSe is the mean square error, k is the number of scans per subject (2 for SLIM dataset and 3 for BNU-3), and n is the number of subjects. The resulting distributions of edge-wise ICC values for the GWM-HFN and GM-GM networks were then statistically compared using a paired t-test.

Finally, to provide a more comprehensive assessment of individual-level stability and uniqueness, as suggested by recent literature^30^, we implemented a connectome identifiability framework. This analysis yields three key metrics: (1) Reliability (µ_intra_): Defined as the mean similarity of an individual’s connectome with their own connectome across two time points. This was calculated by taking the mean of the diagonal elements of the subject-by-subject similarity matrix (Session 1 vs. Session 2). (2) Uniformity (µ_inter_): Defined as the mean similarity of an individual’s connectome with the connectomes of all other individuals. This was calculated by taking the mean of the off-diagonal elements of the similarity matrix. (3) Identifiability Score: Defined as the effect size of the difference between the intra- and inter-individual similarity distributions, calculated as: Identifiability= (µ_intra_−µ_inter_)/s_pooled_, where s_pooled_ is the pooled standard deviation of the two distributions. These three metrics were calculated for both GWM-HFN and GM-GM networks. The Reliability and Uniformity scores were compared between methods using paired t-tests. The statistical significance of the difference in Identifiability scores was assessed using a bootstrap procedure (10,000 iterations).

### 4.5. Topological Analysis of GWM-HFN (SLIM Dataset)

Before analyzing the topological properties of the GWM-HFN, a sparsity-based thresholding procedure was applied to convert the weighted networks into binary representations. Sparsity is defined as the ratio of actual edges to the maximum possible edges in the network. By applying subject-specific thresholds, this procedure ensures a consistent number of edges across participants and sessions under different analytical approaches. Given the absence of a definitive sparsity standard, we adopted a range of [0.10, 0.34] with an interval of 0.01, following the default settings in DPABINet^70^. This specific range was chosen because it ensures that the resulting networks are both sparse enough to minimize spurious connections and sufficiently dense to maintain full network connectivity, allowing for the stable estimation of small-world properties^71,72^. Subsequently, various graph-based network measures, including small-world properties, modularity, and degree, were calculated using the DPABINet toolbox. Detailed formulas and interpretations of these measures are available in the relevant literature^73^. For this analysis, we utilized the SLIM time1 dataset.

#### Small world Organization

Firstly, to determine whether the GWM-HFN exhibits small-world properties, commonly observed in brain networks, we calculated key small-world metrics such as the clustering coefficient and characteristic path length for each subject’s connectivity network. These global measures were normalized by comparing them to the mean values of 100 randomly rewired networks generated using the degree-preserving rewiring algorithm implemented in the Brain Connectivity Toolbox within DPABINet. This algorithm randomizes the network’s topology by iteratively swapping pairs of edges while precisely preserving the number of nodes, edges, and, crucially, the degree distribution of the original network. This ensures that the null models serve as a rigorous benchmark, differing only in their topological organization, not in their basic degree structure. The observed clustering coefficient (C_p_) and characteristic path length (L_p_) of the actual network were then normalized by dividing them by the mean C_p,rand_ and L_p,rand_ from the 100 corresponding random networks. This yields the normalized metrics gamma (γ = C_p_ / C_p,rand_) and lambda (λ = L_p_ / L_p,rand_). A network is considered to exhibit small-world properties if it meets the criteria of γ > 1 and λ ≈ 1, indicating that it is significantly more clustered than a random network but has a similarly efficient path length.

#### Modularity

We also calculated the modularity coefficient Q, which quantifies the degree to which a network can be divided into distinct modules or communities. The Q value is between 0∼1 and the real network modularity function value is generally between 0.3∼0.7^74^. Higher values of Q indicate a more pronounced modular structure, suggesting that the network is composed of tightly connected clusters with relatively sparse connections between them.

#### Degree Distribution and Hubs

To investigate the roles of individual nodes in GWM-HFN, we calculated the nodal degree for GWM-HFN across a range of sparsity thresholds. In the context of brain networks, degree distributions often follow power-law-like distributions, suggesting a scale-free organization^75^. Therefore, after averaging the nodal degree across participants and sparsity levels, we applied different models (power law, exponential, and exponentially truncated power law) to fit the degree distribution of the GWM-HFN. Regions with average nodal degrees in the top 15% were classified as hubs.

### 4.6. Comparison of GWM-HFN and GM-GM Networks (SLIM Dataset)

The GM-GM network captures direct functional interactions between GM regions, whereas the GWM-HFN incorporates WM functional connectivity profiles, thereby representing a more complex and indirect connectivity structure. It is crucial to determine whether the GWM-HFN provides unique information that offers additional value beyond the GM-GM network. To this end, we conducted a series of analyses across multiple dimensions, including edge-level correlations, population variability, and network organization, highlighting their similarities and differences.

#### Similarity Assessment

We first evaluated the similarities between the two networks through Pearson correlation analyses at three levels: edge, network, and global. At the edge level, Pearson correlation coefficients were calculated for each connection, comparing the connectivity values derived from GWM-HFN and GM-GM networks. At the network level, we computed the average intra-network and inter-network connectivity strengths across seven functional subnetworks for both methods and analyzed their correlation. Finally, at the global level, the mean connectivity values across all edges were calculated for each participant, and the correlation coefficients across participants were determined to assess the overall consistency between the two approaches.

#### Variability Assessment

To quantitatively compare the spatial topographies of inter-individual variability between the GWM-HFN and GM-GM frameworks, we implemented the variance decomposition method developed by Mueller et al^33^. This method was applied to our test-retest SLIM dataset to distinguish "true" biological inter-subject variability from session-to-session measurement noise (i.e., intra-subject variability).The model posits that the observed between-subject variability in a given brain node (**Y**) is a linear combination of the true, underlying inter-subject variability (X) and the intra-subject variability, or noise (**N**). By estimating Y and N directly from the test-retest data for each of the 90 brain nodes within each framework, we could derive a residual measure of the true inter-subject variability (**X**) that has been statistically corrected for measurement noise.

Following this decomposition, we conducted three statistical analyses to compare the properties of the true variability maps between the two frameworks: (1) Magnitude Comparison: To test whether the overall magnitude of true inter-individual variability differed between the frameworks, we performed a paired comparison on the 90 node-wise variability values using a non-parametric Wilcoxon signed-rank test. (2) Spatial Similarity Assessment: To quantify the degree of similarity in the spatial patterns of variability, we calculated the Pearson correlation coefficient between the 90-node true variability maps derived from the GWM-HFN and GM-GM frameworks. A 95% confidence interval for the correlation was also estimated. (3) Topographical Overlap Analysis: To further investigate the divergence in the spatial distribution of high-variability regions, we first identified the set of nodes constituting the top 20% most variable regions for each framework separately. We then computed the Jaccard index to quantify the spatial overlap between these two sets of high-variability nodes.

#### Network Organization Analysis

Lastly, we explored the differences in network organization between the two methods. The Jaccard index was employed to measure the structural similarity between GWM-HFN and GM-GM FC networks for each participant across a range of sparsity thresholds (0.01–0.50, with a step size of 0.01). Additionally, we computed global and local graph-theoretical metrics, including multiple global and local indices, using the DPABINet toolbox^70^. Network comparisons were primarily based on paired t-tests performed on the AUC derived from the 0.10-0.34 sparsity range. These metrics provided a detailed comparison of the topological features of the two networks, revealing unique organizational properties attributable to the inclusion of WM functional connectivity profiles in the GWM-HFN.

### 4.7. Age-Related Patterns of GWN-FHN (SALD Dataset)

To investigate age-related patterns and understand how the GWM-HFN evolves over time, we employed linear models to analyze the trajectory of the GWM-HFN throughout adulthood using the SALD dataset. To comprehensively evaluate the unique characteristics of GWM-HFN, we benchmarked our findings against a parallel analysis of traditional GM-GM functional connectivity. The models considered both linear (in Eq. (1)) and quadratic terms (in Eq. (2)) to capture potential non-linear relationships as shown in the following formula, including sex and head motion (measured by mean FD) as covariables,

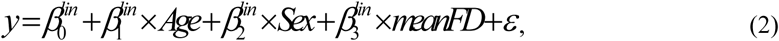

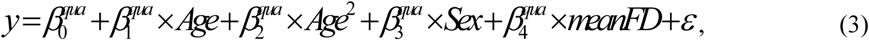

in which dependent variable is GWM-HFN connectivity strength. The model selection was based on the best description of the trajectory of these measures throughout adulthood, determined by the Akaike Information Criterion (AIC). The significance tests of 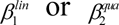 were conducted by t-tests. In the results section, we reported standardized coefficients of 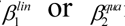 for the best-fit model. Additionally, for models exhibiting a significant quadratic effect of age, the peak age was calculated using the formula for the axis of symmetry in a quadratic function:

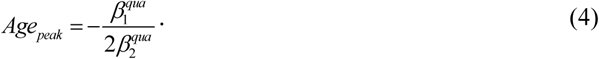

### 4.8. Clinical Correlates of GWM-HFN (ABIDE-II Datasets)

To evaluate the clinical relevance of the GWM-HFN framework, we utilized the ABIDE-II dataset, comparing individuals with ASD against TC. Given the multi-site nature of ABIDE-II, we first applied a harmonization procedure to mitigate potential site-related biases. Specifically, the ComBat algorithm^37^, implemented via the neuroCombat R package (https://github.com/Jfortin1/neuroCombat_Rpackage/), was used to harmonize the raw GM-WM FC values across acquisition sites before constructing the final GWM-HFN matrices for each participant. ComBat is an empirical Bayes method that adjusts for such ‘batch effects’ by modeling and removing site-specific shifts in both the data’s mean (additive effects) and variance (multiplicative effects). Crucially, by including age, sex, and mean FD as biological covariates in the model, ComBat preserves the variance associated with these variables of interest while harmonizing the data across sites. This technique effectively corrected for potential biases in GWM-HFN connectivity. To rigorously confirm the efficacy of the ComBat harmonization, we conducted a comprehensive quantitative validation. We assessed site effects both before and after harmonization using two approaches: 1) A Type III ANOVA was performed on the global mean connectivity (mean Fisher’s z-transformed correlation) with site as a factor, while controlling for age, sex, mean FD, and diagnosis. 2) At the edge level, we employed nested linear models to calculate the proportion of connections showing a significant site effect (*P*_*FDR*_ < 0.05) and to quantify the median effect size of the site variable using partial eta-squared (η^2^).

Subsequent group comparisons between ASD and TC participants were conducted on the harmonized GWM-HFN data. First, we compared the global mean connectivity strength between the two groups using a two-sample t-test, including age, sex, and mean FD as covariates. Second, edge-wise group differences were assessed using two-sample t-tests for each connection within the GWM-HFN, again accounting for age, sex, and mean FD. Significance levels for edge-wise comparisons were determined after applying FDR correction for multiple comparisons across all edges. The anatomical distribution of significantly altered connections was summarized by quantifying their membership within seven established functional subnetworks. To benchmark the sensitivity of GWM-HFN against conventional methods, the identical analysis pipeline—including global mean comparison, edge-wise t-tests with covariate control and FDR correction, and network distribution summary—was repeated using traditional GM-GM functional connectivity networks derived from the same participants.

Finally, we investigated the relationship between GWM-HFN connectivity alterations and clinical symptom severity in the ASD group. Specifically, we calculated the Spearman rank correlation between the mean connectivity strength of the GWM-HFN edges showing significant group differences and the participants’ total scores on the ADOS_G_TOTAL.

### 4.9. Behavioral and Cognitive Correlates of GWM-HFN (BGSP Dataset)

The BGSP dataset included a broad range of behavioral and cognitive measures, which allowed examining phenotypic correlates of GWM-HFN. To provide a direct benchmark, a parallel analysis was conducted on conventional GM-GM networks. Specifically, we used the PLS regression to examine the ability of each connectome to explain interindividual variance in 4 behavioral and cognitive phenotypes, including shipley vocabulary score, matrix reasoning scores, shipley and matrix reasoning estimated IQ. In the PLS regression model, the response variable was behavioral and cognitive data for a certain domain and the predictor variables were all edges in the respective connectome. Only the first component of the PLS model (i.e., PLS1) was examined, which was the linear combination of GWM-HFN connections that exhibited the strongest correlation with the behavioral and cognitive data. Significance levels of the correlations were estimated by randomly shuffling the behavioral and cognitive data among participants (10,000 times). For each significant correlation, the contribution of a given edge was defined as its weight to form the PLS1. Furthermore, to ensure the robustness of these findings, we employed a bootstrap procedure (with 10,000 resamples) to assess the statistical stability of the PLS feature weights.

For measures that showed significant associations in the preceding PLS analyses, we further employed the BBS modeling method^38^ to evaluate the predictive capacity of both the GWM-HFN and the conventional GM-GM networks for individual scores. The same analytical pipeline was applied independently to each network type. First, principal component analysis was used for dimensionality reduction by retaining components that explained 80% variance in interregional morphological similarity. Then, a linear regression model was fitted between the expression scores of the retained components and behavioral and cognitive data, which was further used to predict behavioral and cognitive outcomes for unseen participants. Finally, the Pearson correlation between actual scores and predicted values was calculated for each behavioral and cognitive domain. To assess the performance of the BBS model, a ten-fold cross-validation procedure was used. Since a single cross-validation might be sensitive to a particular split of the data into folds^76^, the ten-fold cross-validation procedure was repeated 100 times and the resulting mean Pearson correlation coefficient was reported for each behavioral and cognitive domain. To test whether the Pearson correlation coefficients were significantly higher than random operations, a nonparametric permutation testing procedure was performed by reshuffling the behavioral and cognitive data and repeating the ten-fold cross-validation procedure (10000 times). For each significant correlation, the contribution of a given edge was calculated as the mean value of the product of the coefficient in principal component analysis with the beta value in the linear regression model across all folds and repetitions. For the visualization of these contributing connectivity patterns, we selected edges with absolute standardized weights exceeding 3 standard deviations (|z| > 3σ). This stringent threshold was chosen to ensure that only the most robustly contributing edges were displayed, thereby enhancing the interpretability of the underlying neurobiological pattern.

### 4.10. Statistics and Reproducibility

All statistical analyses were performed using custom scripts in combination with publicly available toolboxes, including SPM12 (http://www.fil.ion.ucl.ac.uk/spm/software/spm12), DPARSF (http://rfmri.org/DPARSF), and DPABINet, and the neuroCombat R package. Unless otherwise specified, a two-tailed P-value < 0.05 was considered statistically significant. For analyses involving multiple comparisons across network edges or nodes, the FDR was used to correct P-values, with a significance threshold of P_FDR_ <0.05.

The topological properties of the GWM-HFN, such as small-worldness, were statistically validated by comparing network metrics against an ensemble of 100 null networks generated using a degree-preserving rewiring algorithm. Test-retest reliability was assessed at multiple levels: edge-wise reliability was quantified using the Intraclass Correlation Coefficient (ICC(2,1)), and connectome-level stability and uniqueness were evaluated using an identifiability framework, with statistical significance of differences between frameworks (GWM-HFN vs. GM-GM) assessed via paired t-tests or bootstrap procedures (10,000 iterations).

Inter-individual variability was compared between frameworks using the Mueller et al. variance decomposition method to separate true biological variance from measurement noise, with statistical comparisons performed using non-parametric Wilcoxon signed-rank tests, Pearson correlation, and the Jaccard index. Age-related trajectories were modeled using linear and quadratic regression, controlling for sex and head motion, with model selection based on the Akaike Information Criterion. Group comparisons in the clinical analysis (ASD vs. TC) were conducted using two-sample t-tests on ComBat-harmonized data, controlling for age, sex, and mean FD. The efficacy of harmonization was quantitatively validated using Type III ANOVA and partial eta-squared effect sizes.

Brain-behavior relationships were assessed using two multivariate approaches. Partial least squares regression was used to identify connectivity patterns associated with cognitive domains, with significance determined by permutation testing (10,000 permutations) and feature stability assessed via bootstrapping (10,000 resamples). The predictive utility of these patterns for individual scores was then validated using a brain bias set modeling approach within a repeated (100 times) ten-fold cross-validation framework, with prediction significance also assessed via permutation testing (10,000 permutations).

Reproducibility of the findings was ensured through rigorous validation across six large-scale, independent, and publicly available datasets (SLIM, BNU-3, SALD, ABIDE-II, BGSP, and the validation analysis on the Schaefer atlas with the SLIM dataset), with specific sample sizes for each analysis detailed in the relevant sections and Supplementary Table S3. The use of both short-term (BNU-3 dataset) and long-term (SLIM dataset) test-retest data provided a robust assessment of the reliability of the proposed GWM-HFN framework.

## Ethical Statement

All procedures performed in studies involving human participants were in accordance with the ethical standards of the institutional and/or national research committee and with the 1964 Helsinki declaration and its later amendments or comparable ethical standards. This article does not contain any studies with animals performed by any of the authors. Informed consent was obtained from all individual participants included in the study.

## Data Availability

All MRI data used in this study were derived from publicly available resources, including: the Southwest University Longitudinal Imaging Multimodal (SLIM) dataset, available at http://dx.doi.org/10.15387/fcp_indi.retro.slim; the Beijing Normal University (BNU-3) dataset, available at http://dx.doi.org/10.15387/fcp_indi.corr.bnu3; the Southwest University Adult Lifespan (SALD) dataset, available at https://fcon_1000.projects.nitrc.org/indi/retro/sald.html; the Autism Brain Imaging Data Exchange II (ABIDE-II) project, available at https://fcon_1000.projects.nitrc.org/indi/abide/abide_II.html; and the Brain Genomics Superstruct Project (BGSP) dataset, available at https://www.neuroinfo.org/gsp/. The numerical source data for graphs and charts presented in the main figures are available in the Supplementary Data file.

## Code Availability

The custom code used for the GWM-HFN construction and subsequent statistical analyses in this study will be made publicly available on GitHub upon acceptance of the manuscript. The analyses were performed using custom scripts in combination with publicly available toolboxes, including SPM12 (http://www.fil.ion.ucl.ac.uk/spm/software/spm12), DPARSF (http://rfmri.org/DPARSF), and DPABINet.

## Authors Contribution

Z.L. conceived and designed the overall study. Z.L. and K.H. oversaw data collection and verified the analytical methods. Z.L., K.H., Z.C., Y.X. and Y.C. performed data preprocessing, constructed the networks, and carried out the statistical analyses. Z.L., and K.H. contributed to the methodology design and drafted large parts of the manuscript. Z.L., K.H., and C.Z. provided critical revisions, shaped the discussion of results, and coordinated manuscript editing. All authors discussed the results, commented on the manuscript at all stages, and approved the final version for publication.

## Conflict of Interest Statement

The authors declare that they have no conflict of interest.

## Financial support

Kang Hu is supported by Wuhan Business School Doctoral Fund Project (2024KB016), and MOE-LCSM. Ze-Qiang Linli is supported by the Humanities and Social Sciences Youth Foundation of Ministry of Education of China (24YJCZH164), Guangdong Province General University Characteristic Innovation projects (2024KTSCX178).

## Notes

### Competing Interest Statement

The authors have declared no competing interest.

### Funding Statement

Zeqiang Linli is supported by the Humanities and Social Sciences Youth Foundation of Ministry of Education of China (24YJCZH164),and the Guangdong Province General University Characteristic Innovation projects (2024KTSCX178).

### Author Declarations

Publicly available datasets were analyzed in this study.

### Summary of Updates

revision according the reviewer's comments from the submmitted Journal

