## Supporting Information for "GWM-HFN: A Novel Gray-White Matter Heterogeneous Fusion Network for Functional Connectomes"

### Study Datasets

#### ABIDE-II

The Autism Brain Imaging Data Exchange II (ABIDE-II) is a large-scale, open-access neuroimaging database established to advance research on the brain connectome in Autism Spectrum Disorder (ASD) (https://fcon_1000.projects.nitrc.org/indi/abide/abide_II.html). Building on the success of ABIDE-I, ABIDE-II addresses the substantial heterogeneity of ASD and the complexity of the connectome by aggregating more extensive and better-characterized datasets. It comprises contributions from 19 international sites, including 10 founding institutions and 7 new members, and includes a total of 1,114 datasets (521 ASD individuals and 593 typically developing controls (TC)) spanning a wide age range (5–64 years). The datasets incorporate structural MRI (sMRI), resting-state functional MRI (rs-fMRI), and detailed phenotypic information, particularly regarding core ASD traits and associated symptoms.

In this study, participants with missing functional imaging data, poor image quality, or excessive head motion were excluded. Furthermore, sites with fewer than 25 participants were also excluded to ensure robust statistical power. As a result, the final dataset included 640 participants (272 ASD and 368 TC) from 13 sites.

#### BGSP

The Brain Genomics Superstruct Project (BGSP) is a comprehensive neuroimaging dataset that integrates imaging, behavioral, cognitive, and personality data, providing an invaluable resource for studying brain function and structure in healthy individuals (https://www.neuroinfo.org/gsp). This large-scale dataset includes over 1,500 participants and captures a wide range of variables such as age, BMI, and gender. The neuroimaging data consist of high-quality sMRI scans and rs-fMRI scans. In this study, following rigorous quality control procedures, a total of 1,564 participants were included in the final analyses.

#### BNU-3

The Beijing Normal University 3 (BNU-3) dataset is part of Consortium for Reliability and Reproducibility (CoRR) project, including 48 healthy controls from a community (student) sample from Beijing Normal University in China. Each participant has 3 rs-fMRI scans (https://fcon_1000.projects.nitrc.org/indi/CoRR/html/bnu_3.html). During the first scan participants were instructed to rest with their eyes closed. The second and third resting state scan were randomized between resting with eyes open versus eyes closed. In this study, a total of 46 participants were included in the final analysis. The dataset was primarily utilized to assess the short-term test-retest reliability of the gray-white matter heterogeneous fusion network (GWM-HFN).

#### SALD

The Southwest University Adult Lifespan (SLAD) dataset, which encompasses a substantial cross-sectional sample of 494 healthy controls, ranging in age from 19 to 80 years, who underwent a comprehensive multi-modal neuroimaging examination, including sMRI and rs-fMRI (http://fcon_1000.projects.nitrc.org/indi/retro/sald.html). In this study, following rigorous quality control procedures, a total of 440 participants were included in the final analyses.

#### SLIM

The Southwest University Longitudinal Imaging Multimodal (SLIM) dataset is a longitudinal neuroimaging dataset that includes data from healthy young adults, predominantly university students. The dataset features multimodal MRI data, including structural, diffusion-weighted, and resting-state functional MRI, collected at three time points over a span of approximately 3.5 years (https://fcon_1000.projects.nitrc.org/indi/retro/southwestuni_qiu_index.html). At the time point 1, the average age of all the subjects is 20.08 (range 17 – 27) and the median is 20. There are 323 females, 257 males (time point 1) and 121 females, 119 males (time point 2), and 135 females, 93 males (time point 3) participated in the scanning. In this study, we primarily used time1 and time2 data to construct the GWM-HFN and evaluate its long-term test-retest (TRT) reliability. The average number of days between the first scan and the second scan are 304.14 days. In our study, we included 572 participants at time1 and 237 participants at time2, with 219 participants being consistent across both time points.

#### Mapping AAL atlas to Macroscopic Networks

The localization of nodes and clusters was identified using the Automated Anatomical Labeling (AAL) atlas[1]. Additionally, we applied the "winner-takes-all" approach to map the AAL brain regions onto Yeo's 7-network model[2]. Detailed correspondence between regions and networks can be found in Table S1 for further reference.

We initially aligned each Region of Interest (ROI) from the AAL-90 brain template with the Yeo-7 network template. Subsequently, we examined the proportion of the seven components of the Yeo network within each ROI and categorized each ROI into the network with the highest overlapping proportion, adhering to the principle of "winner-takes-all". Subsequently, we made adjustments based on previous research and anatomical considerations. The primary focus of these adjustments involved reclassifying regions that were clearly misclassified and addressing areas (e.g., subcortex regions) that could not be categorized due to minimal overlap between the AAL and Yeo templates.

The core region of the Visual Network (VN) lies within the occipital lobe. Combining the researches conducted by Smith et al.[3] and Weiner et al.[4], along with the anatomical positions within the Yeo network for the VN, we have identified a total of 14 nodes. The core regions of the Sensorimotor Network (SMN) are located within the motor and somatosensory cortices. Combining prior research and the anatomical positions within the Yeo network, we have identified a total of 14 nodes within the SMN[2,5]. In conjunction with the study conducted by Jimenez et al.[6], we have identified six nodes within the ventral attention network. Drawing upon the studies conducted by Uddin et al.[7], Vossel et al.[8], and Jimenez et al.[6], we have identified four nodes within the dorsal attention network. In this study, these ventral attention network and dorsal attention network nodes together constitute the Attention Network (AN), which therefore comprises a total of 10 nodes. Combining Catani et al[9].'s comprehensive review of the limbic system and the anatomical positions within Yeo's limbic network, we have identified 16 nodes within the limbic network (LN). Drawing upon the research conducted by Oliver et al.[10], anatomical observations within the Yeo network, and insights provided by Uddin et al[7]., we have ultimately identified 10 nodes within the Frontoparietal Network (FPN). Based on the anatomical locations of the Default Mode Network (DMN) within Yeo's 7-network model[2] and the core nodes of the DMN provided by Uddin et al.[7], in conjunction with previous researches on the default network[11,12], a total of 18 nodes were identified. The Basal Ganglia Network (BGN) were determined based on their anatomical locations, which consists of 8 ROIs.

### Supplementary Tables

**Table S1. Names, abbreviations, and functional network affiliations of the 90 AAL regions**

| AAL Order | AAL Label | Abbreviation | Network Name |
|---|---|---|---|
| 1 | Precentral_L | PreCG.L | Sensorimotor Network |
| 2 | Precentral_R | PreCG.R | Sensorimotor Network |
| 3 | Frontal_Sup_L | SFGdor.L | Dorsal Attention Network |
| 4 | Frontal_Sup_R | SFGdor.R | Dorsal Attention Network |
| 5 | Frontal_Sup_Orb_L | ORBsup.L | Limbic Network |
| 6 | Frontal_Sup_Orb_R | ORBsup.R | Limbic Network |
| 7 | Frontal_Mid_L | MFG.L | Frontoparietal Network |
| 8 | Frontal_Mid_R | MFG.R | Frontoparietal Network |
| 9 | Frontal_Mid_Orb_L | ORBmid.L | Frontoparietal Network |
| 10 | Frontal_Mid_Orb_R | ORBmid.R | Frontoparietal Network |
| 11 | Frontal_Inf_Oper_L | IFGoperc.L | Frontoparietal Network |
| 12 | Frontal_Inf_Oper_R | IFGoperc.R | Frontoparietal Network |
| 13 | Frontal_Inf_Tri_L | IFGtriang.L | Frontoparietal Network |
| 14 | Frontal_Inf_Tri_R | IFGtriang.R | Frontoparietal Network |
| 15 | Frontal_Inf_Orb_L | ORBinf.L | Default Mode Network |
| 16 | Frontal_Inf_Orb_R | ORBinf.R | Default Mode Network |
| 17 | Rolandic_Oper_L | ROL.L | Sensorimotor Network |
| 18 | Rolandic_Oper_R | ROL.R | Sensorimotor Network |
| 19 | Supp_Motor_Area_L | SMA.L | Sensorimotor Network |
| 20 | Supp_Motor_Area_R | SMA.R | Sensorimotor Network |
| 21 | Olfactory_L | OLF.L | Limbic Network |
| 22 | Olfactory_R | OLF.R | Limbic Network |
| 23 | Frontal_Sup_Medial_L | SFGmed.L | Default Mode Network |
| 24 | Frontal_Sup_Medial_R | SFGmed.R | Default Mode Network |
| 25 | Frontal_Med_Orb_L | ORBsupmed.L | Default Mode Network |
| 26 | Frontal_Med_Orb_R | ORBsupmed.R | Default Mode Network |
| 27 | Rectus_L | REC.L | Limbic Network |
| 28 | Rectus_R | REC.R | Limbic Network |
| 29 | Insula_L | INS.L | Ventral Attention Network |
| 30 | Insula_R | INS.R | Ventral Attention Network |
| 31 | Cingulum_Ant_L | ACG.L | Default Mode Network |
| 32 | Cingulum_Ant_R | ACG.R | Default Mode Network |
| 33 | Cingulum_Mid_L | DCG.L | Ventral Attention Network |
| 34 | Cingulum_Mid_R | DCG.R | Ventral Attention Network |
| 35 | Cingulum_Post_L | PCG.L | Default Mode Network |
| 36 | Cingulum_Post_R | PCG.R | Default Mode Network |
| 37 | Hippocampus_L | HIP.L | Limbic Network |

| | | | |
|---|---|---|---|
| 38 | Hippocampus_R | HIP.R | Limbic Network |
| 39 | ParaHippocampal_L | PHG.L | Default Mode Network |
| 40 | ParaHippocampal_R | PHG.R | Default Mode Network |
| 41 | Amygdala_L | AMYG.L | Limbic Network |
| 42 | Amygdala_R | AMYG.R | Limbic Network |
| 43 | Calcarine_L | CAL.L | Visual Network |
| 44 | Calcarine_R | CAL.R | Visual Network |
| 45 | Cuneus_L | CUN.L | Visual Network |
| 46 | Cuneus_R | CUN.R | Visual Network |
| 47 | Lingual_L | LING.L | Visual Network |
| 48 | Lingual_R | LING.R | Visual Network |
| 49 | Occipital_Sup_L | SOG.L | Visual Network |
| 50 | Occipital_Sup_R | SOG.R | Visual Network |
| 51 | Occipital_Mid_L | MOG.L | Visual Network |
| 52 | Occipital_Mid_R | MOG.R | Visual Network |
| 53 | Occipital_Inf_L | IOG.L | Visual Network |
| 54 | Occipital_Inf_R | IOG.R | Visual Network |
| 55 | Fusiform_L | FFG.L | Visual Network |
| 56 | Fusiform_R | FFG.R | Visual Network |
| 57 | Postcentral_L | PoCG.L | Sensorimotor Network |
| 58 | Postcentral_R | PoCG.R | Sensorimotor Network |
| 59 | Parietal_Sup_L | SPG.L | Dorsal Attention Network |
| 60 | Parietal_Sup_R | SPG.R | Dorsal Attention Network |
| 61 | Parietal_Inf_L | IPL.L | Frontoparietal Network |
| 62 | Parietal_Inf_R | IPL.R | Frontoparietal Network |
| 63 | SupraMarginal_L | SMG.L | Ventral Attention Network |
| 64 | SupraMarginal_R | SMG.R | Ventral Attention Network |
| 65 | Angular_L | ANG.L | Default Mode Network |
| 66 | Angular_R | ANG.R | Default Mode Network |
| 67 | Precuneus_L | PCUN.L | Default Mode Network |
| 68 | Precuneus_R | PCUN.R | Default Mode Network |
| 69 | Paracentral_Lobule_L | PCL.L | Sensorimotor Network |
| 70 | Paracentral_Lobule_R | PCL.R | Sensorimotor Network |
| 71 | Caudate_L | CAU.L | Basal Ganglia Network |
| 72 | Caudate_R | CAU.R | Basal Ganglia Network |
| 73 | Putamen_L | PUT.L | Basal Ganglia Network |
| 74 | Putamen_R | PUT.R | Basal Ganglia Network |
| 75 | Pallidum_L | PAL.L | Basal Ganglia Network |
| 76 | Pallidum_R | PAL.R | Basal Ganglia Network |
| 77 | Thalamus_L | THA.L | Basal Ganglia Network |
| 78 | Thalamus_R | THA.R | Basal Ganglia Network |
| 79 | Heschl_L | HES.L | Sensorimotor Network |
| 80 | Heschl_R | HES.R | Sensorimotor Network |
| 81 | Temporal_Sup_L | STG.L | Sensorimotor Network |

| | | | |
|---|---|---|---|
| 82 | Temporal_Sup_R | STG.R | Sensorimotor Network |
| 83 | Temporal_Pole_Sup_L | TPOsup.L | Limbic Network |
| 84 | Temporal_Pole_Sup_R | TPOsup.R | Limbic Network |
| 85 | Temporal_Mid_L | MTG.L | Default Mode Network |
| 86 | Temporal_Mid_R | MTG.R | Default Mode Network |
| 87 | Temporal_Pole_Mid_L | TPOmid.L | Limbic Network |
| 88 | Temporal_Pole_Mid_R | TPOmid.R | Limbic Network |
| 89 | Temporal_Inf_L | ITG.L | Limbic Network |
| 90 | Temporal_Inf_R | ITG.R | Limbic Network |

**Table S2. Labels of JHU-ICBM- WM 48 Atlas**

| Order | Label |
|---|---|
| 1 | Middle cerebellar peduncle |
| 2 | Pontine crossing tract (a part of MCP) |
| 3 | Genu of corpus callosum |
| 4 | Body of corpus callosum |
| 5 | Splenium of corpus callosum |
| 6 | Fornix (column and body of fornix) |
| 7 | Corticospinal tract R |
| 8 | Corticospinal tract L |
| 9 | Medial lemniscus R |
| 10 | Medial lemniscus L |
| 11 | Inferior cerebellar peduncle R |
| 12 | Inferior cerebellar peduncle L |
| 13 | Superior cerebellar peduncle R |
| 14 | Superior cerebellar peduncle L |
| 15 | Cerebral peduncle R |
| 16 | Cerebral peduncle L |
| 17 | Anterior limb of internal capsule R |
| 18 | Anterior limb of internal capsule L |
| 19 | Posterior limb of internal capsule R |
| 20 | Posterior limb of internal capsule L |
| 21 | Retrolenticular part of internal capsule R |
| 22 | Retrolenticular part of internal capsule L |
| 23 | Anterior corona radiata R |
| 24 | Anterior corona radiata L |
| 25 | Superior corona radiata R |
| 26 | Superior corona radiata L |
| 27 | Posterior corona radiata R |
| 28 | Posterior corona radiata L |
| 29 | Posterior thalamic radiation (include optic radiation) R |
| 30 | Posterior thalamic radiation (include optic radiation) L |
| 31 | Sagittal stratum (include inferior longitidinal fasciculus and inferior fronto-occipital fasciculus) R |
| 32 | Sagittal stratum (include inferior longitidinal fasciculus and inferior fronto-occipital fasciculus) L |
| 33 | External capsule R |
| 34 | External capsule L |
| 35 | Cingulum (cingulate gyrus) R |
| 36 | Cingulum (cingulate gyrus) L |
| 37 | Cingulum (hippocampus) R |
| 38 | Cingulum (hippocampus) L |
| 39 | Fornix (cres) / Stria terminalis (can not be resolved with current resolution) R |
| 40 | Fornix (cres) / Stria terminalis (can not be resolved with current resolution) L |
| 41 | Superior longitudinal fasciculus R |
| 42 | Superior longitudinal fasciculus L |
| 43 | Superior fronto-occipital fasciculus (could be a part of anterior internal capsule) R |
| 44 | Superior fronto-occipital fasciculus (could be a part of anterior internal capsule) L |
| 45 | Uncinate fasciculus R |
| 46 | Uncinate fasciculus L |
| 47 | Tapetum R |
| 48 | Tapetum L |

#### Table S3. Demographic variables of all participants in each dataset

| Dataset | N | Sex (F/M) | Age[a)] (years) | mean FD |
|---|---|---|---|---|
| ABIDE-II | 640 | 138/502 | 15.62±10.15 | 0.10±0.06 |
| ASD | 272 | 39/233 | 15.44±10.48 | 0.11±0.06 |
| TC | 368 | 99/269 | 15.75±9.90 | 0.09±0.05 |
| BGSP | 1564 | 904/660 | 21.54±2.89 | 0.06±0.03 |
| BNU-3 (CoRR) | 46 | 24/22 | 22.50±2.19 | |
| SALD | 440 | 277/163 | 43.57±17.20 | 0.09±0.04 |
| SLIM | | | | |
| Time1 | 572 | 20.08±1.27 | 318/252 | 0.06±0.03 |
| Time2 | 237 | 20.82±1.17 | 120/116 | 0.06±0.02 |

F, female; M, male;

SLIM, Southwest University Longitudinal Imaging Multimodal datasets; BNU-3, BNU 3 - Beijing Normal University (Zang) from Consortium for Reliability and Reproducibility (CoRR) project; SALD, Southwest University Adult Lifespan Dataset, NYU (ADHD-200), New York University from Attention Deficit Hyperactivity Disorder 200 project; CHCP, Chinese Human Connectome Project.

**Table S4. Comparison of Hub Regions between GWM-HFN and GM-GM**

| ID | Full Name | Abbr. | Network | Hub Type | Degree (GWM-HFN) | Degree (GMGM) |
|---|---|---|---|---|---|---|
| 33 | Cingulum_Mid_L | DCG.L | Ventral Attention Network | Common Hub | 30.63 | 34.31 |
| 34 | Cingulum_Mid_R | DCG.R | Ventral Attention Network | Common Hub | 29.22 | 31.07 |
| 47 | Lingual_L | LING.L | Visual Network | Common Hub | 25.88 | 28.39 |
| 48 | Lingual_R | LING.R | Visual Network | Common Hub | 26.56 | 30.79 |
| 51 | Occipital_Mid_L | MOG.L | Visual Network | Common Hub | 28.36 | 32.56 |
| 55 | Fusiform_L | FFG.L | Visual Network | Common Hub | 25.91 | 30.62 |
| 82 | Temporal_Sup_R | STG.R | Sensorimotor Network | Common Hub | 30.33 | 28.18 |
| 85 | Temporal_Mid_L | MTG.L | Default Mode Network | Common Hub | 31.07 | 30.76 |
| 86 | Temporal_Mid_R | MTG.R | Default Mode Network | Common Hub | 30.71 | 31.39 |
| 89 | Temporal_Inf_L | ITG.L | Limbic Network | Common Hub | 25.81 | 29.04 |
| 90 | Temporal_Inf_R | ITG.R | Limbic Network | Common Hub | 31.45 | 35.28 |
| 16 | Frontal_Inf_Orb_R | ORBinf.R | Default Mode Network | GWM-HFN-specific | 25.64 | 22.49 |
| 81 | Temporal_Sup_L | STG.L | Sensorimotor Network | GWM-HFN-specific | 26.35 | 23.88 |
| 84 | Temporal_Pole_Sup_R | TPOsup.R | Limbic Network | GWM-HFN-specific | 27.50 | 26.14 |
| 4 | Frontal_Sup_R | SFGdor.R | Dorsal Attention Network | GM-GM-specific | 25.54 | 27.34 |
| 43 | Calcarine_L | CAL.L | Visual Network | GM-GM-specific | 23.17 | 26.22 |
| 56 | Fusiform_R | FFG.R | Visual Network | GM-GM-specific | 24.77 | 29.70 |

### Supplementary Figures

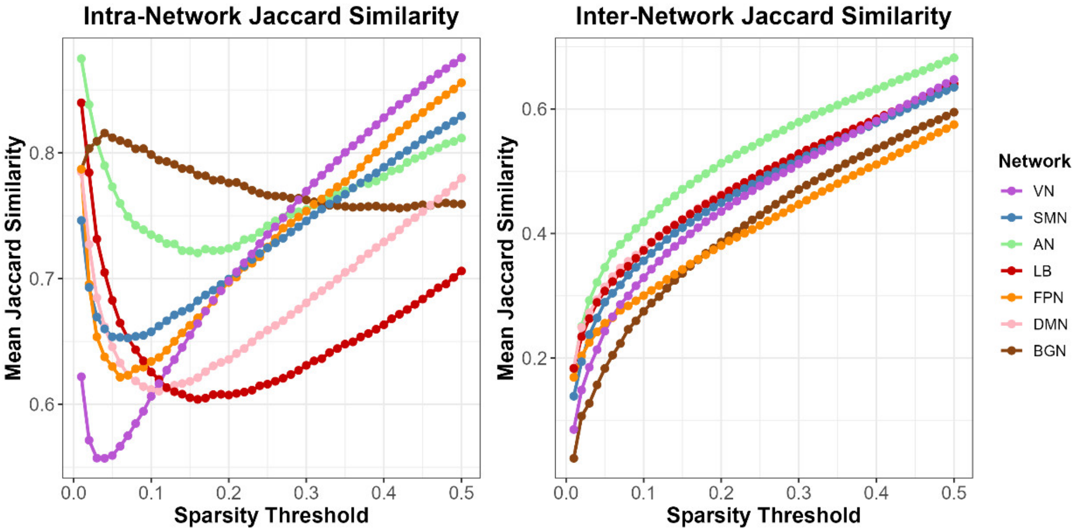

**Figure S1.** Similarity of network-level connectivity between GWM-HFN and GM-GM methods across varying sparsity thresholds. The left panel illustrates the Jaccard coefficient similarity of intra-network connections within respective functional subnetworks as a function of sparsity. The right panel shows the Jaccard coefficient similarity of inter-network connections between different functional subnetworks as a function of sparsity. This figure elucidates the structural differences and the evolution of similarity at the functional module level between the two network construction approaches (GWM-HFN and GM-GM) under different network densities.

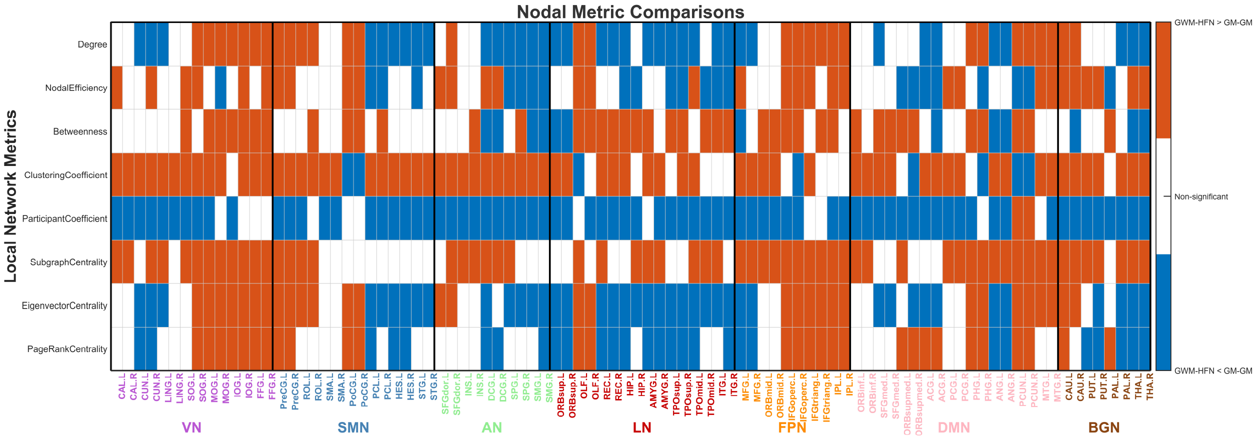

**Figure S2.** Comparison of nodal network attributes between GWM-HFN and GM-GM methods. This figure presents a comparison of various nodal network properties, calculated using the area under the curve (AUC) over a sparsity range of 0.10–0.34, between GWM-HFN and GM-GM networks. The figure clearly displays the reorganized topological characteristics at the nodal level in GWM-HFN networks, emphasizing the unique contribution of incorporating white matter pathway information in functional network analysis.

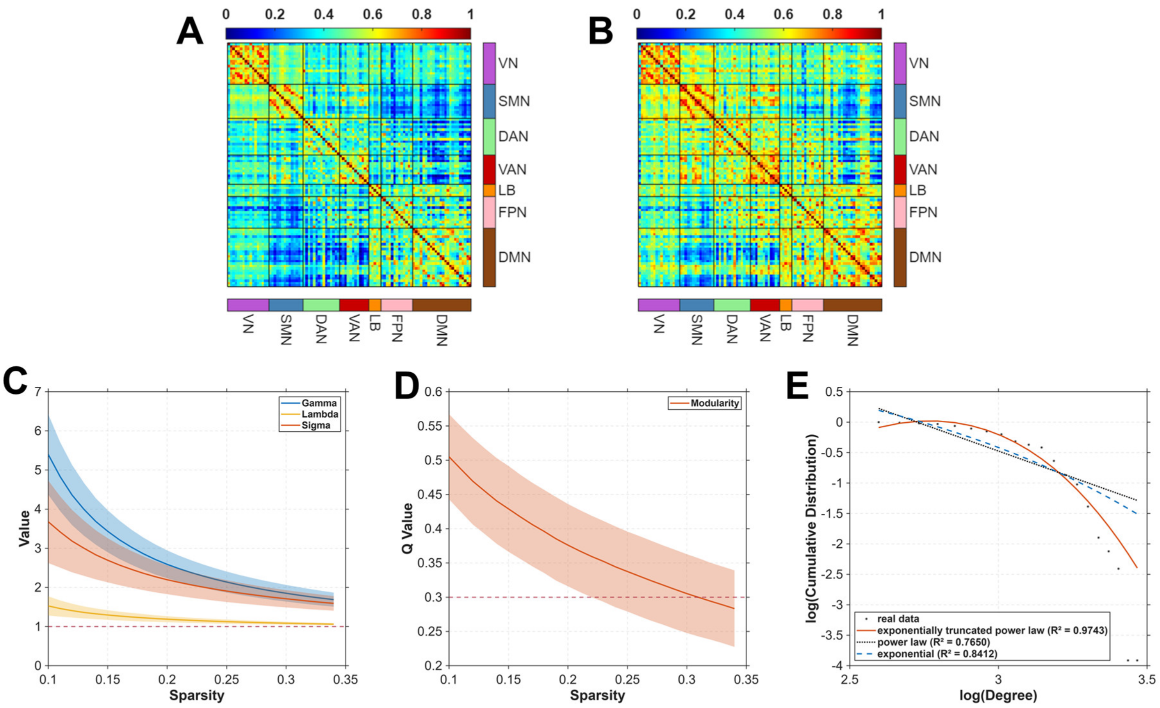

**Figure S3. Validation of GWM-HFN Foundational Properties Using the Schaefer-100 Parcellation.** (A) The group-level mean GM-GM functional connectivity matrix constructed using the Schaefer-100 atlas. The matrix, organized by the Yeo 7-network parcellation, shows strong intra-network connectivity, particularly within the Visual (VN), Somatomotor (SMN), and Default Mode (DMN) networks. (B) The group-level mean GWM-HFN connectivity matrix, also constructed using the Schaefer-100 atlas. It similarly exhibits robust intra-network connectivity but also shows qualitatively stronger inter-network connectivity compared to the conventional GM-GM network. (C) Small-world properties—sigma (σ), normalized clustering coefficient (γ), and normalized characteristic path length (λ)—of the Schaefer-100 based GWM-HFN across a range of sparsity thresholds, presented as mean ± standard deviation. (D) Modularity coefficient (Q) of the Schaefer-100 based GWM-HFN across varying sparsity thresholds, indicating robust modular organization. (E) Degree distribution of the group-averaged, Schaefer-100 based GWM-HFN, which is best fitted by an exponentially truncated power-law model, revealing the existence of highly connected brain regions.
